# Top 50 Cited Articles on Pediatric Respiratory Infections: A Web of Science Bibliometric Analysis

**DOI:** 10.64898/2026.05.18.26353534

**Authors:** Shadan A. Albakri, Ghadeer S. Almasoudi, Deem A. Albakri, Jomana F. Aljariry, Layan B. Aljohny, Leena N. Rizg, Lujain M. Alzahrani, Elaaf A. Albadi, Latifa A. Alsubaie, Wajd B. Alyoubi, Abeer A. Alnajjar

## Abstract

**Background:** Pediatric respiratory infections are a leading cause of morbidity and mortality globally, representing a major health challenge in children.

**Research Gap:** Despite extensive studies on epidemiology, clinical management, and specific pathogens, no bibliometric analysis has systematically evaluated the most influential research in this field.

**Objectives:** This study aimed to evaluate the characteristics of the top 50 most-cited articles on pediatric respiratory infections and to identify emerging research trends.

**Methods:** The Web of Science database was searched without publication year restrictions. Independent reviewers screened studies based on predefined inclusion and exclusion criteria. Data were extracted using a standardized form, including study details.

**Results:** The 50 most-cited articles ranged from 34 to 384 citations and showed a right-skewed distribution with a sharp drop after the top ten. Publication years ranged from 1978 to 2021, with over half published in the 2010s. Articles appeared in 31 journals, with ***Pediatrics*** contributing five. Leading countries were the United States (18%), China (12%), and Canada (10%), with research largely concentrated in high-income regions and limited multicenter collaboration. Cohort studies dominated (66%), while randomized trials (12%) and reviews/meta-analyses (16%) were less common. Research clustered around three themes: clinical outcomes (e.g., pneumonia, bronchiolitis); viral etiology/diagnostics (e.g., RSV, SARS-CoV-2); and antimicrobial stewardship.

**Conclusion:** Over the past decades, pediatric respiratory infection research has developed but remains unbalanced, relying heavily on observational evidence from high-income countries, with limited randomized trials, systematic reviews, multicenter collaborations, and LMIC-led studies. These findings provide insights that may direct researchers to identify potential focal points and guide future research in the field.

## Introduction

Pediatric respiratory tract infections are a major global health problem and leading cause of child morbidity and mortality [1]. Viruses such as RSV, influenza, and parainfluenza increase hospitalization and mortality, particularly in children under five [2]. Diagnosis is challenging because respiratory viral infections present with overlapping clinical features, making laboratory confirmation essential for accurate identification [3]. Early accurate diagnosis is essential to guide appropriate treatment, prevent unnecessary antibiotic use and reduce complications [4].

Bacterial co-infections, including influenza-associated bacteria and Streptococcus pneumoniae, may complicate and worsen RSV infections severity [5]. Additionally, children with underlying conditions or immunocompromised status are at higher risk of complications, including lung transplant recipients, showing a significantly reduced one-year survival [6].

Despite advances in prevention and management, challenges remain. The role of children with SARS-CoV-2 in transmission dynamics and screening strategies is still unclear [7]. Nosocomial RSV infection is associated with prolonged hospitalization and increased morbidity and mortality in high-risk children [8]. Preventive interventions, including probiotics, show only modest benefits in reducing the incidence and severity of upper respiratory tract infections [9]. These persistent challenges highlight the need to identify and critically assess studies with the greatest impact on the field. Although pediatric respiratory research has expanded substantially, no bibliometric analysis has systematically evaluated the most influential studies in this area.

This gap is further compounded by uneven global surveillance, as most data originate from developed countries, with limited representation from developing regions. This lack of consolidated evidence creates a limitation in the understanding of global epidemiology of pediatric respiratory infections [10].

While many studies address specific pathogens and treatment strategies, widely cited articles rarely assess the collective citation impact, methodological quality, or research design, making it difficult to identify studies that have shaped practice, policy, or research priorities [10, 11].

Bibliometric analyses in several pediatric subspecialties have identified influential articles, collaboration networks, and novel developments [11]. However, no similar analysis exists for pediatric respiratory infections. Data limitations are most pronounced in low- and middle-income countries, where disease burden is highest but surveillance remains limited [12,13]. This absence of representation restricts global understanding and hinders region-specific prevention and treatment strategies, underscoring the need for equitable, evidence-based approaches.

## Methods

### 1. Search Strategy

The Web of Science database was used to conduct a comprehensive literature search using targeted keywords such as “Pediatric” and “Respiratory Infections.” The search was not restricted by year of publication to allow broad identification of relevant studies. Only English-language publications specifically focused on pediatric respiratory infections were included.

Although the initial search yielded many articles, particular emphasis was placed on studies with high citation counts, as this reflects their scientific impact and relevance. The identified studies were subsequently organized according to their maximum citation count.

### 2. Study Selection

The literature screening process was conducted by four independent reviewers. Titles and abstracts of retrieved studies were screened for relevance based on predefined inclusion and exclusion criteria. Studies were eligible if they addressed pediatric respiratory infections, included affected patient populations, were published in English, and had been cited in academic literature. A fifth reviewer resolved any disagreements during the screening phase. Exclusion criteria included non-English publications, studies not primarily focused on pediatric respiratory infections, articles from non–peer-reviewed sources, and publications lacking citation records.

### 3. Data Extraction and Management

Data extraction was conducted by four independent reviewers using a standardized form to ensure consistency. Variables included: first author, study type, year of publication, total and average annual citation. Additional data included nutritional strategy, sample size, mean patient age, study setting, country, journal name, impact factor (IF), level of evidence, and primary outcomes. Disagreements were resolved through discussion, with a fifth reviewer consulted if needed.

From 17,416 Web of Science records -which were ranked by total citation count- the top 200 articles were screened in two stages, resulting in the exclusion of 115 studies due to language, lack of focus, non–peer-reviewed sources, or missing citation records. The 50 most-cited articles were included in the final analysis, with 35 additional studies excluded for not ranking in the top 50. This data selection and extraction process is illustrated in Figure 1.

**Figure 1:**
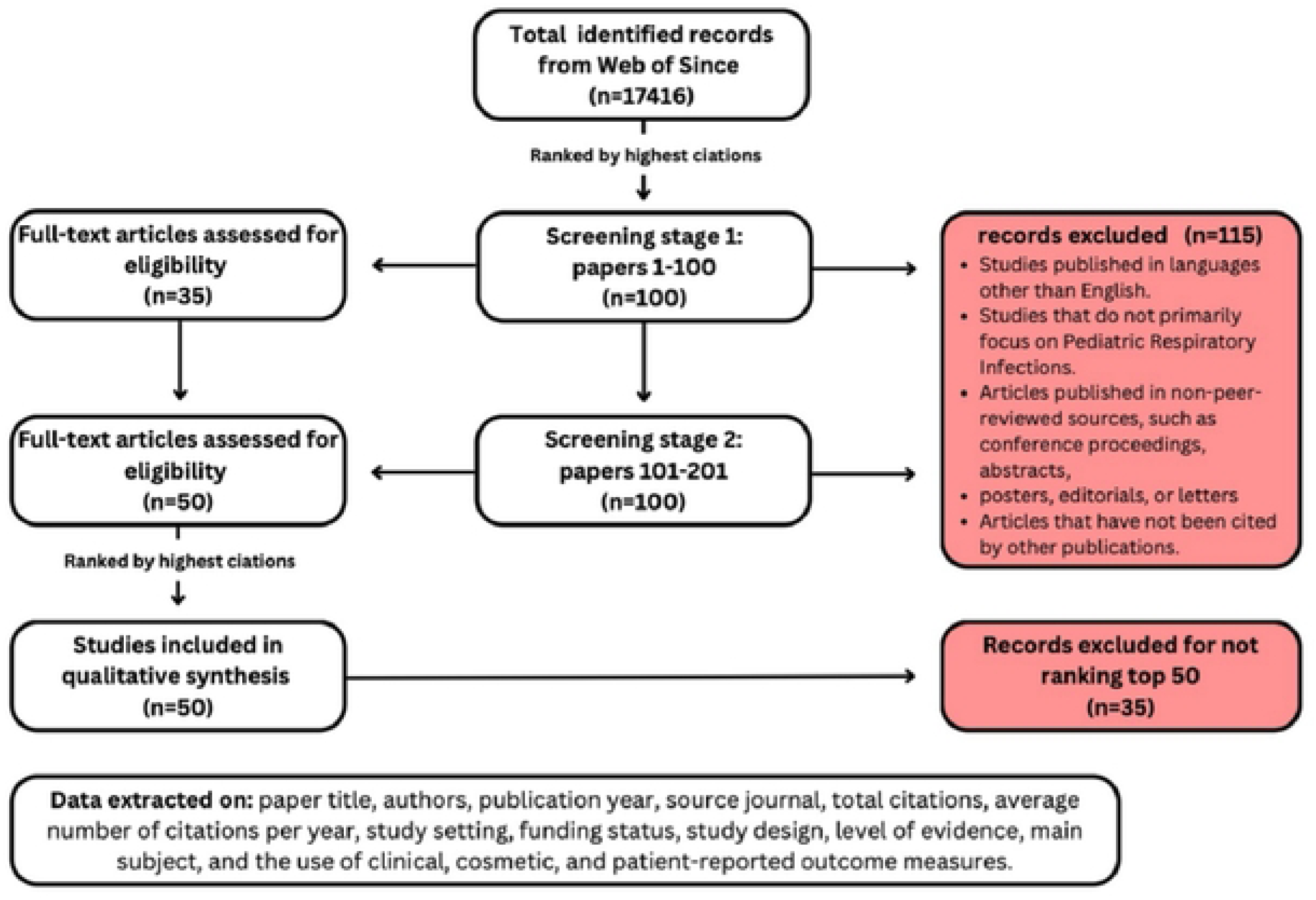
PRISMA chart for screening process.

### 4. Treatment of Missing Data

When data were missing or incomplete, corresponding authors were contacted when possible, to obtain additional information. If authors could not be reached or did not respond, missing data were documented, and their potential impact was addressed in the final analysis.

### 5. Statistical Analysis

Descriptive statistics were used to summarize study characteristics, including means, medians, and ranges for continuous variables such as patient age and citation counts. Categorical variables were summarized using frequency distributions. Citation trends over time were analyzed, and although no formal bias assessment was performed, a qualitative evaluation of methodological rigor and relevance was conducted.

## Results

The 50 most-cited pediatric respiratory infection studies show a strongly right-skewed citation distribution, ranging from 384 to 34 citations, with a sharp decline after the top ten papers (ranks 1–10 = 384–110 vs. ranks 11–20 = 107–68). This indicates a compact cluster of landmark studies that capture much of the field’s attention, as shown in Table 1. Regarding recency, 54% (27/50) were published between 2010 and 2024, reflecting sustained and contemporary citation influence.

**Table 1:**
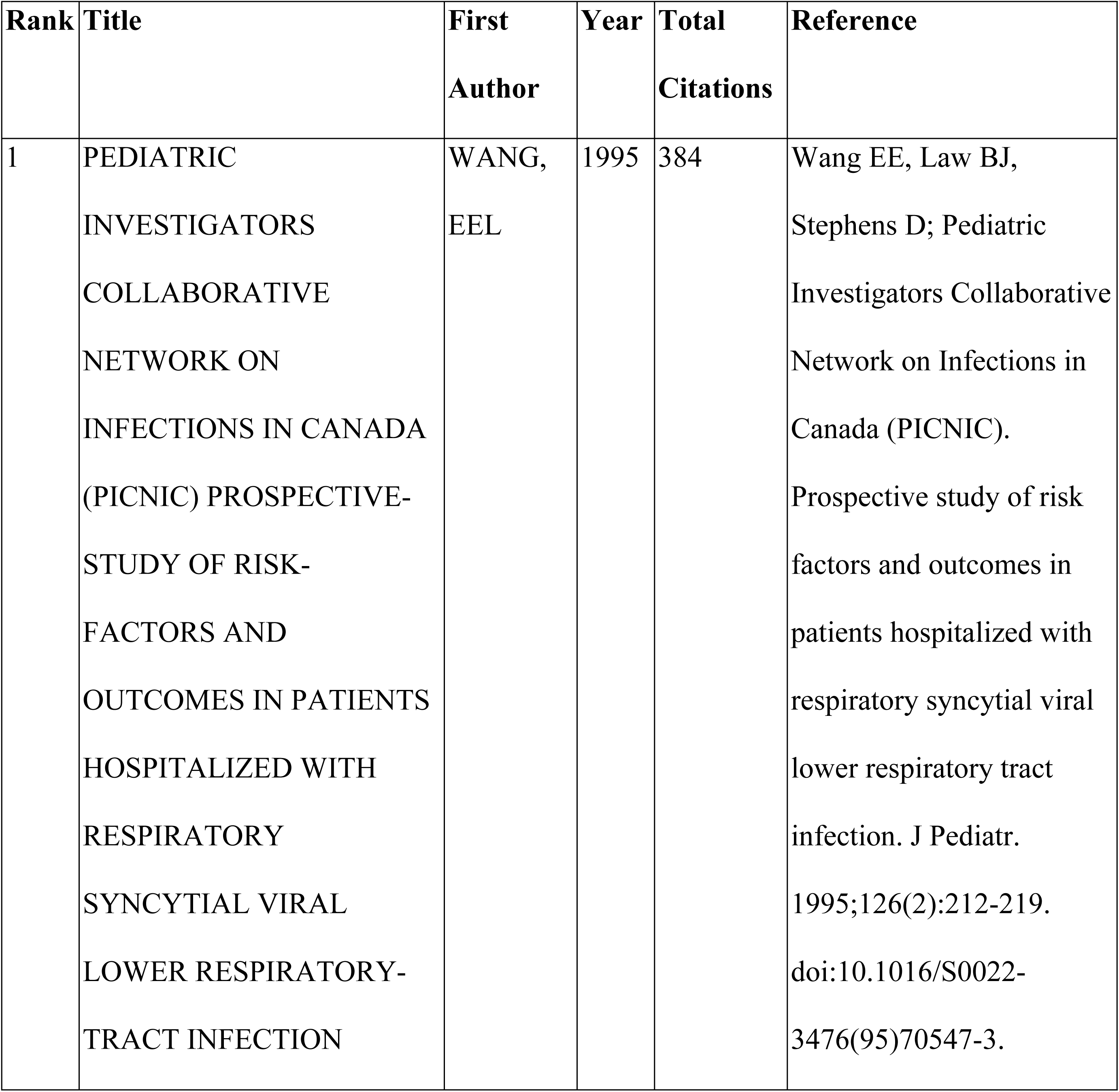
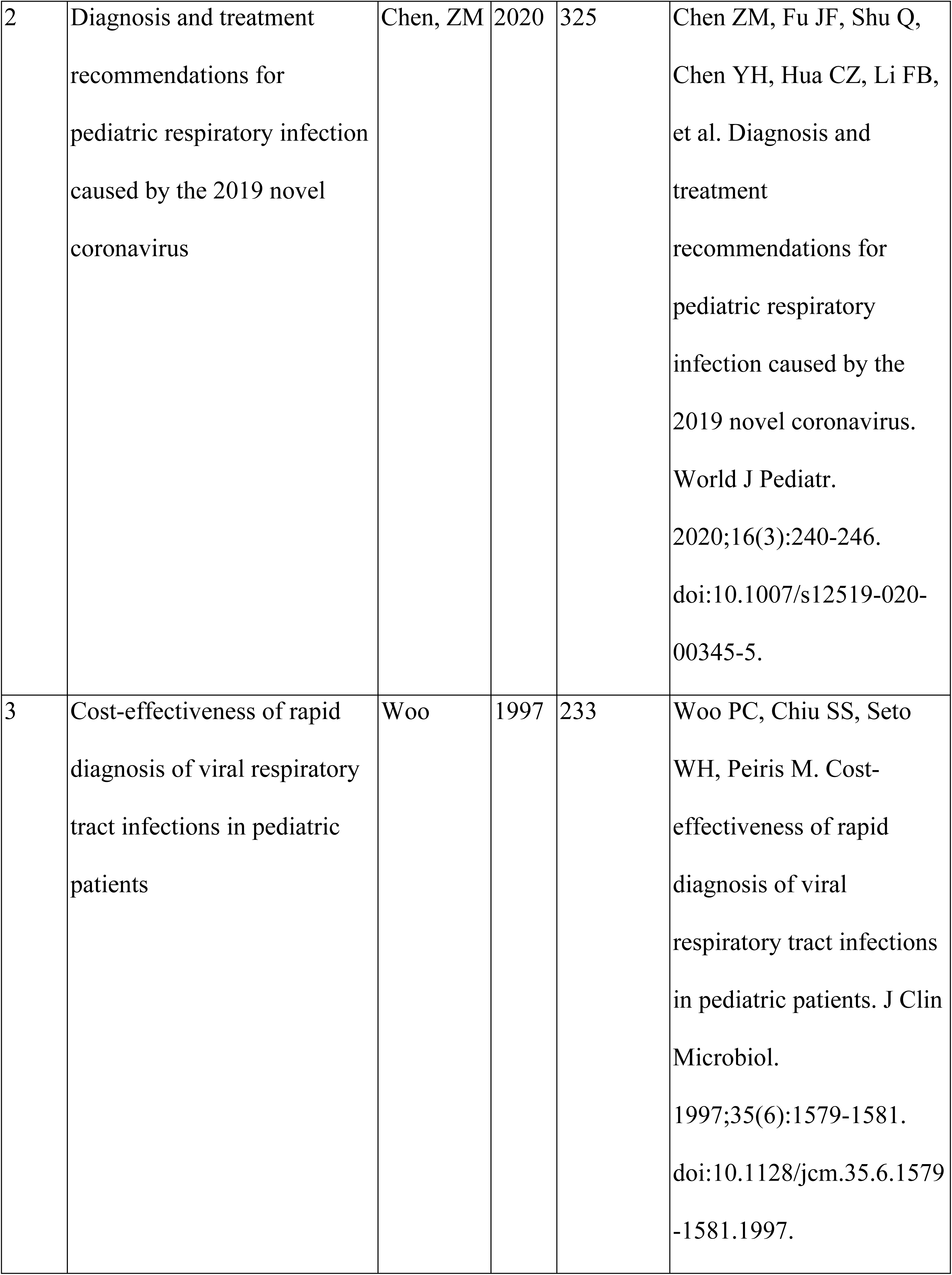
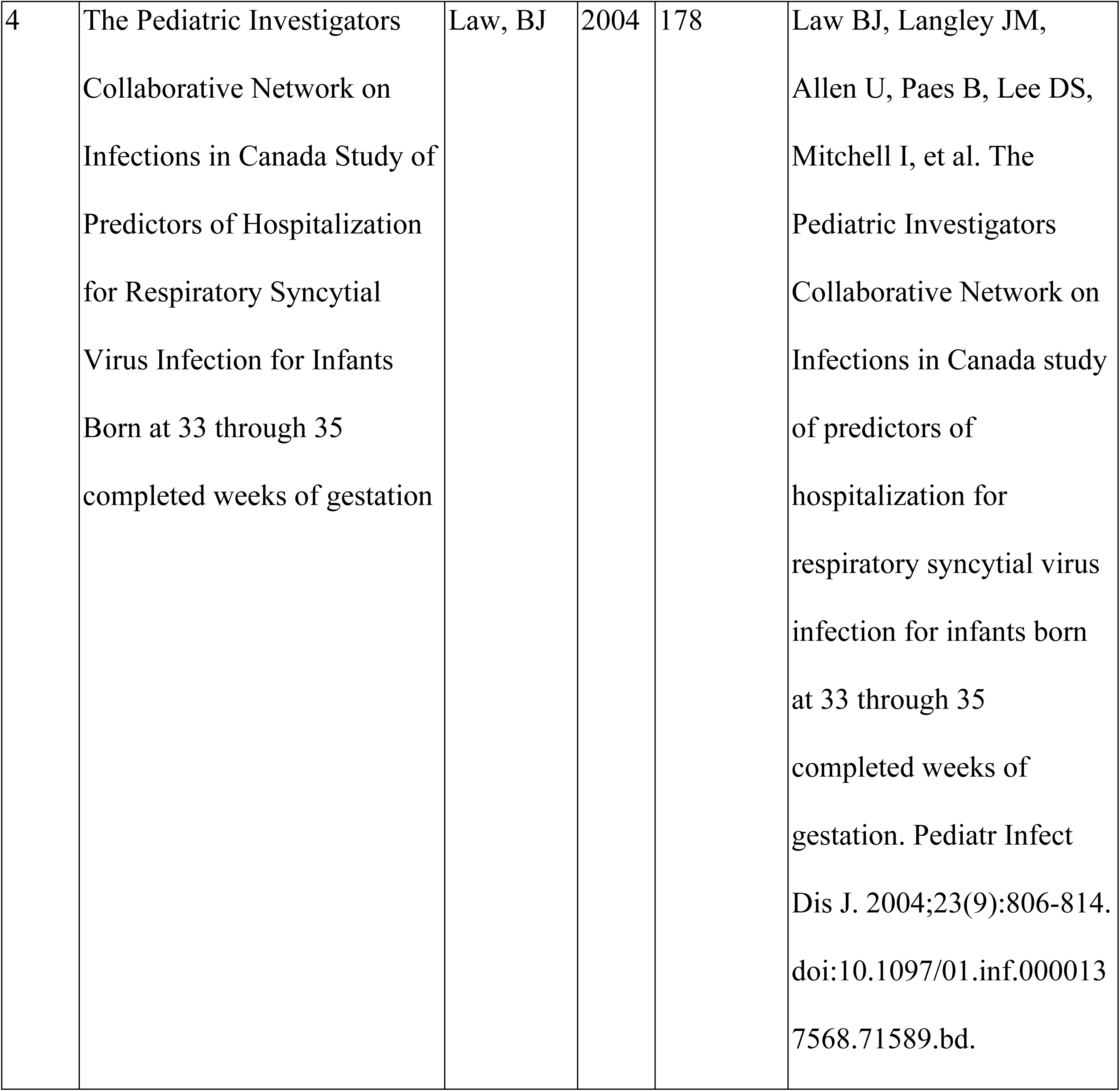
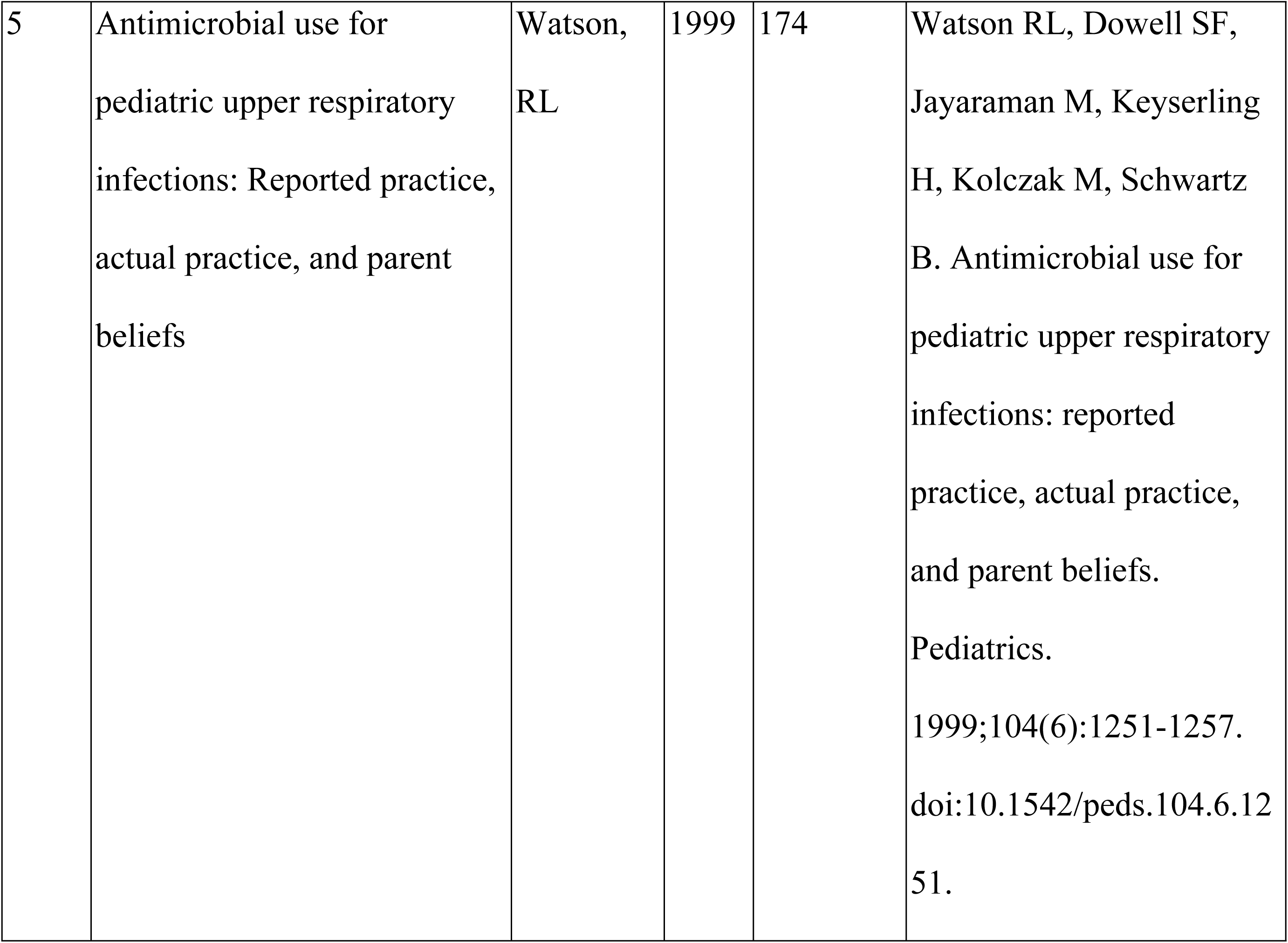
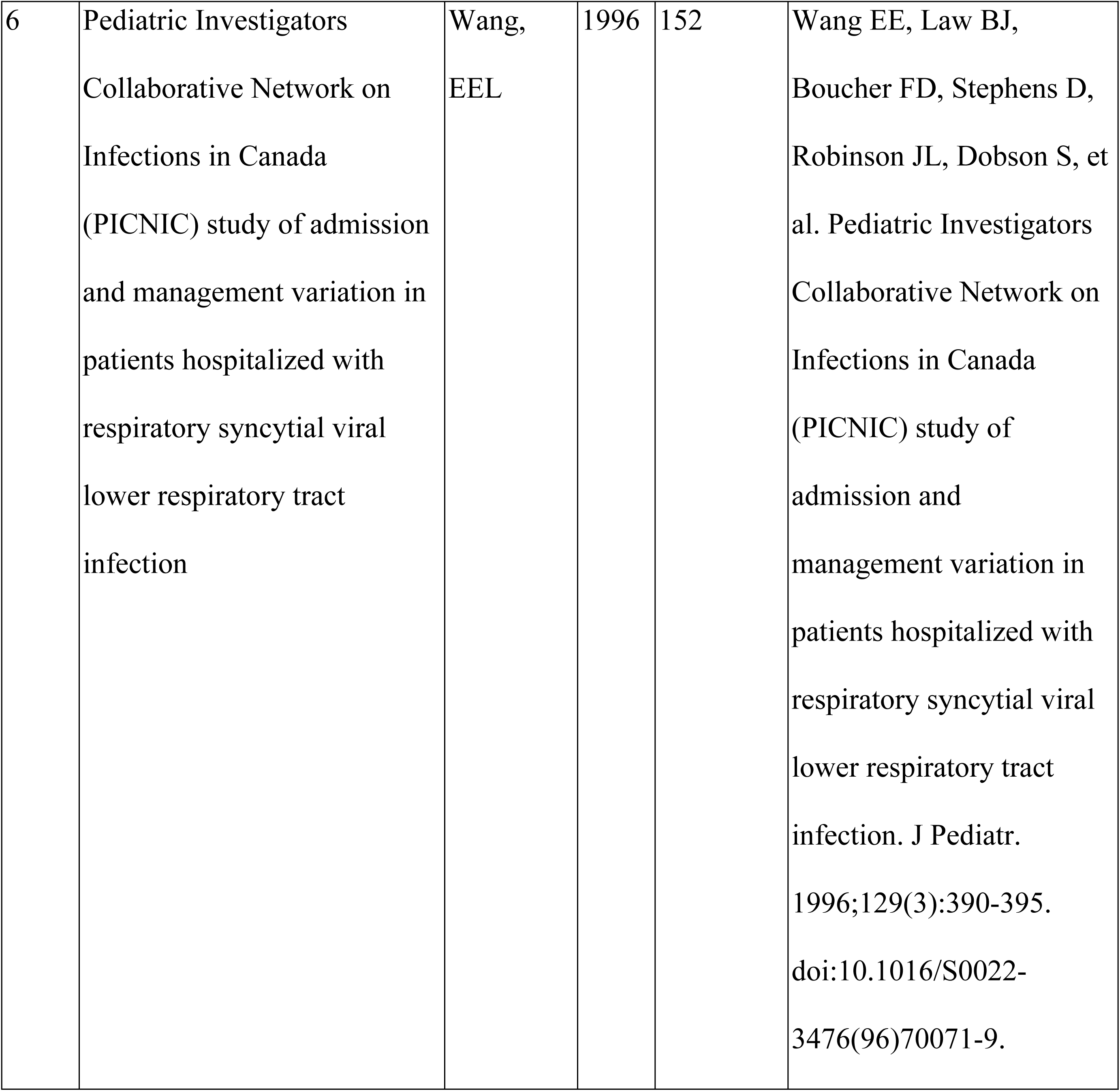
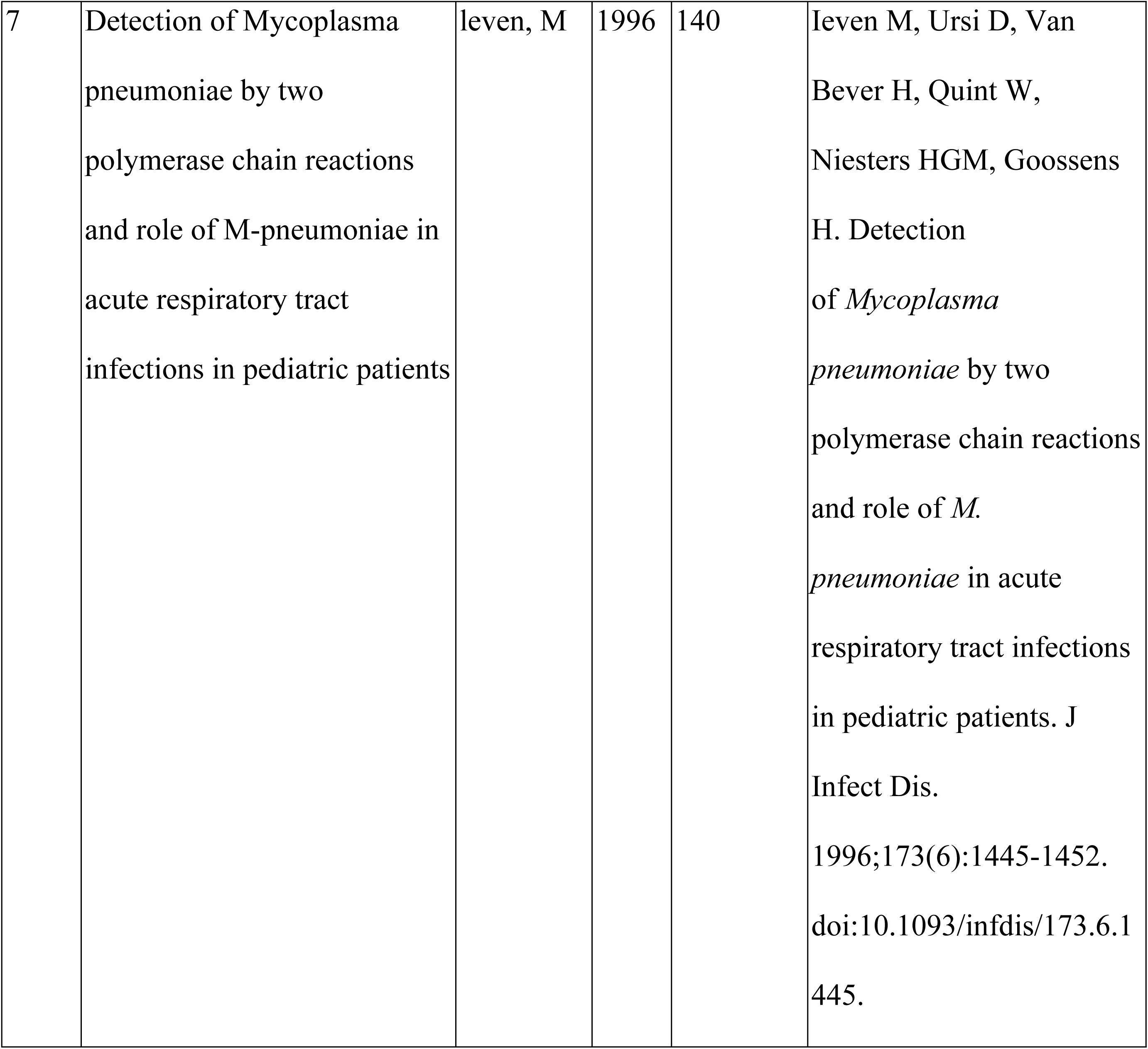
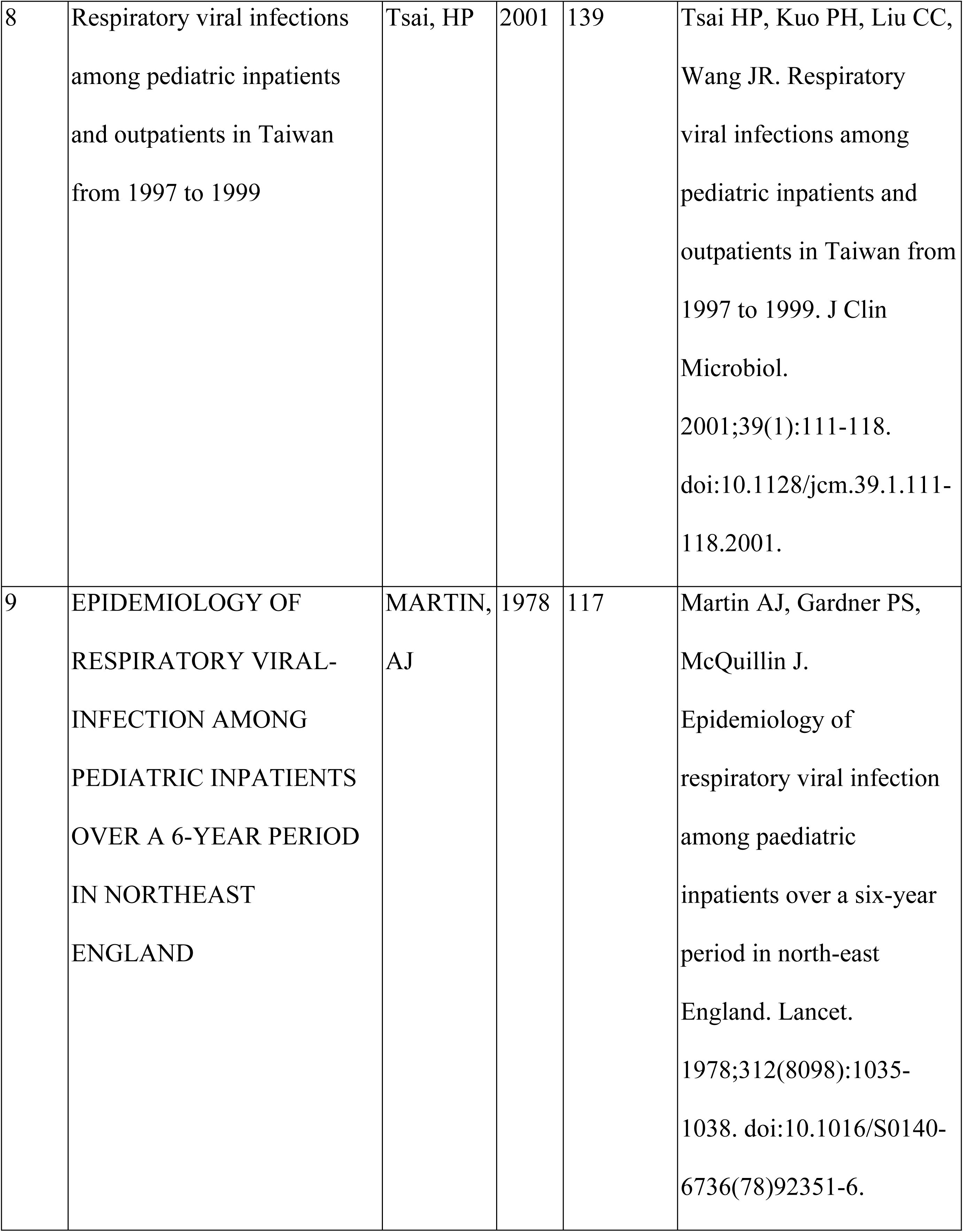
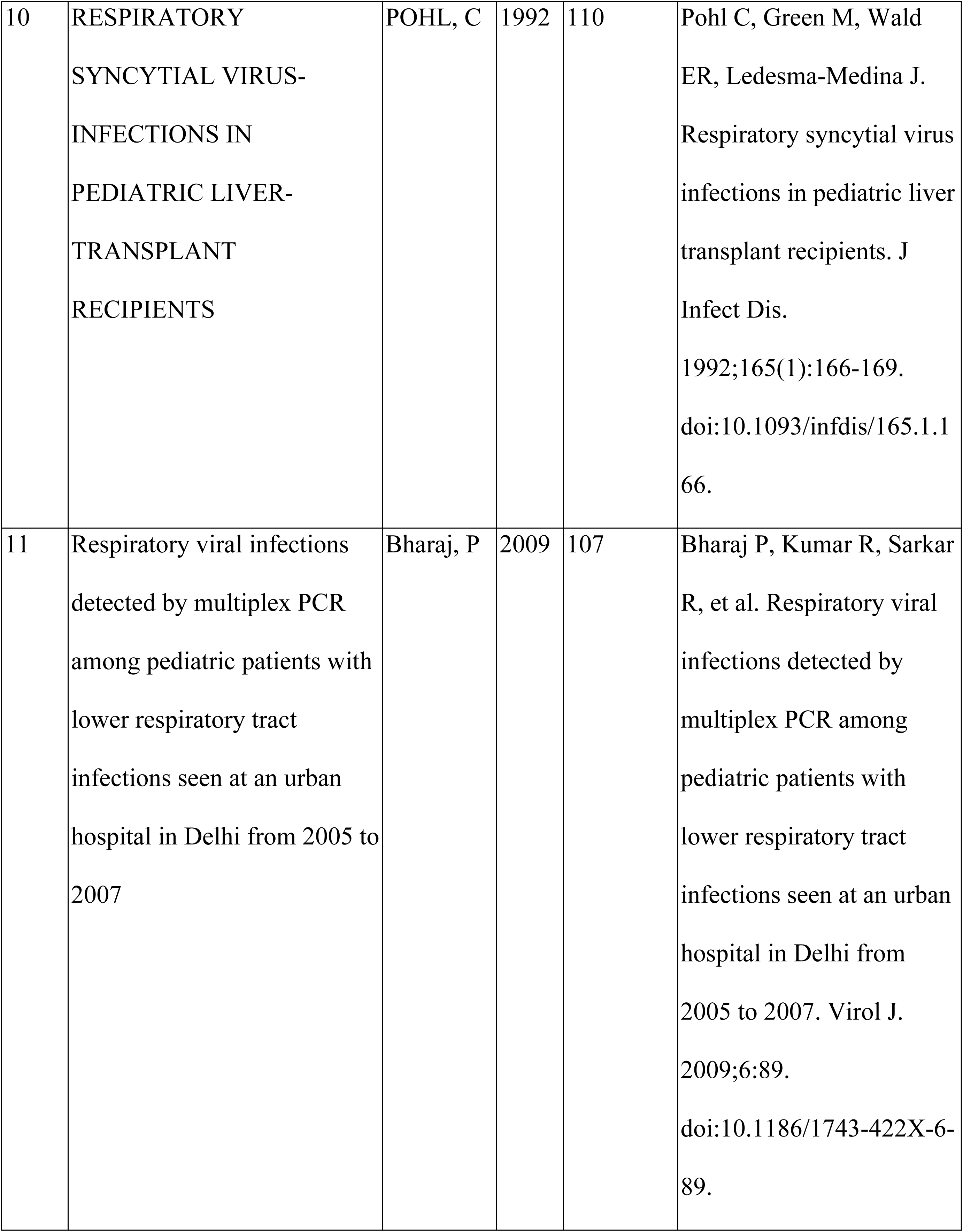
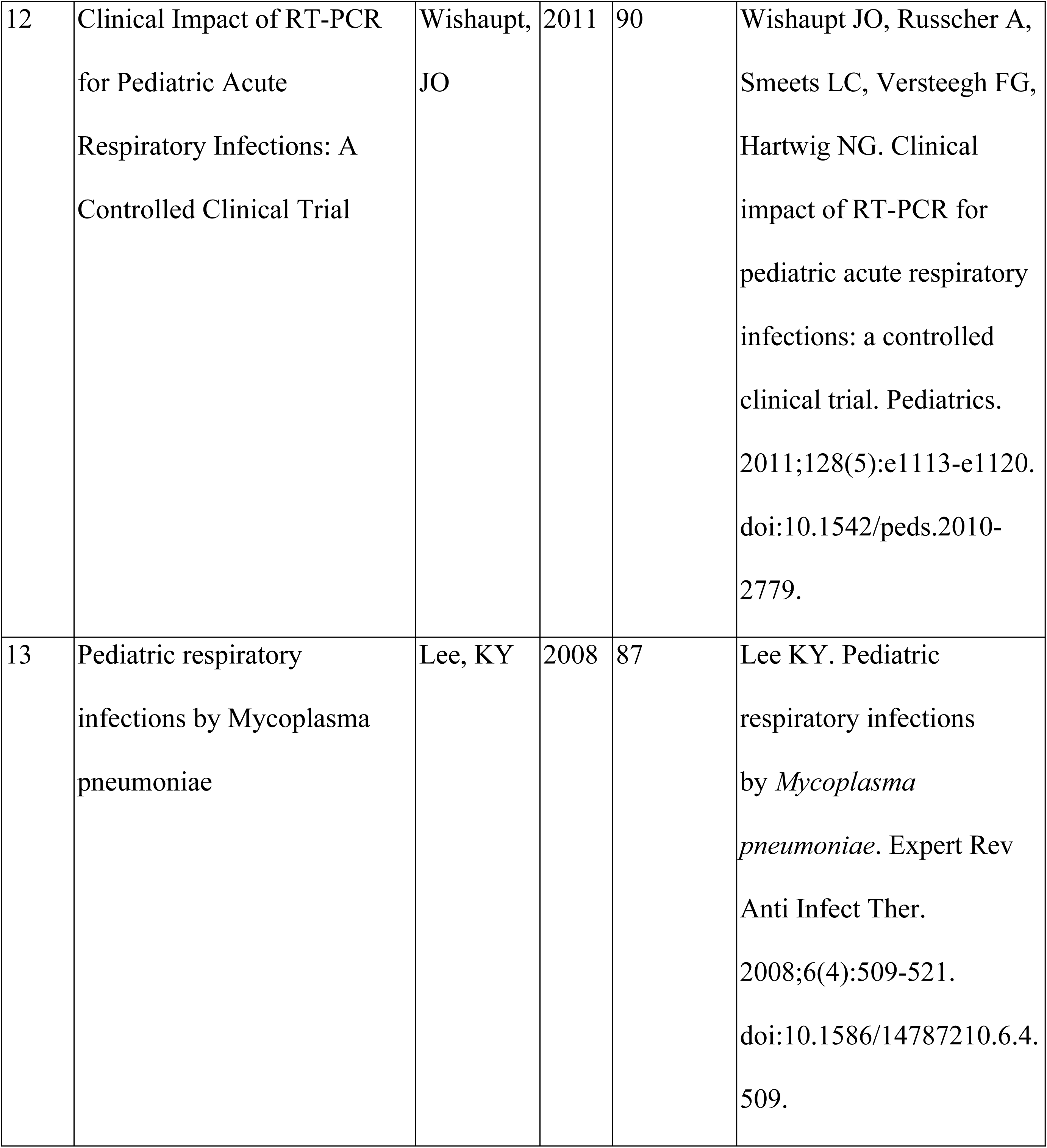
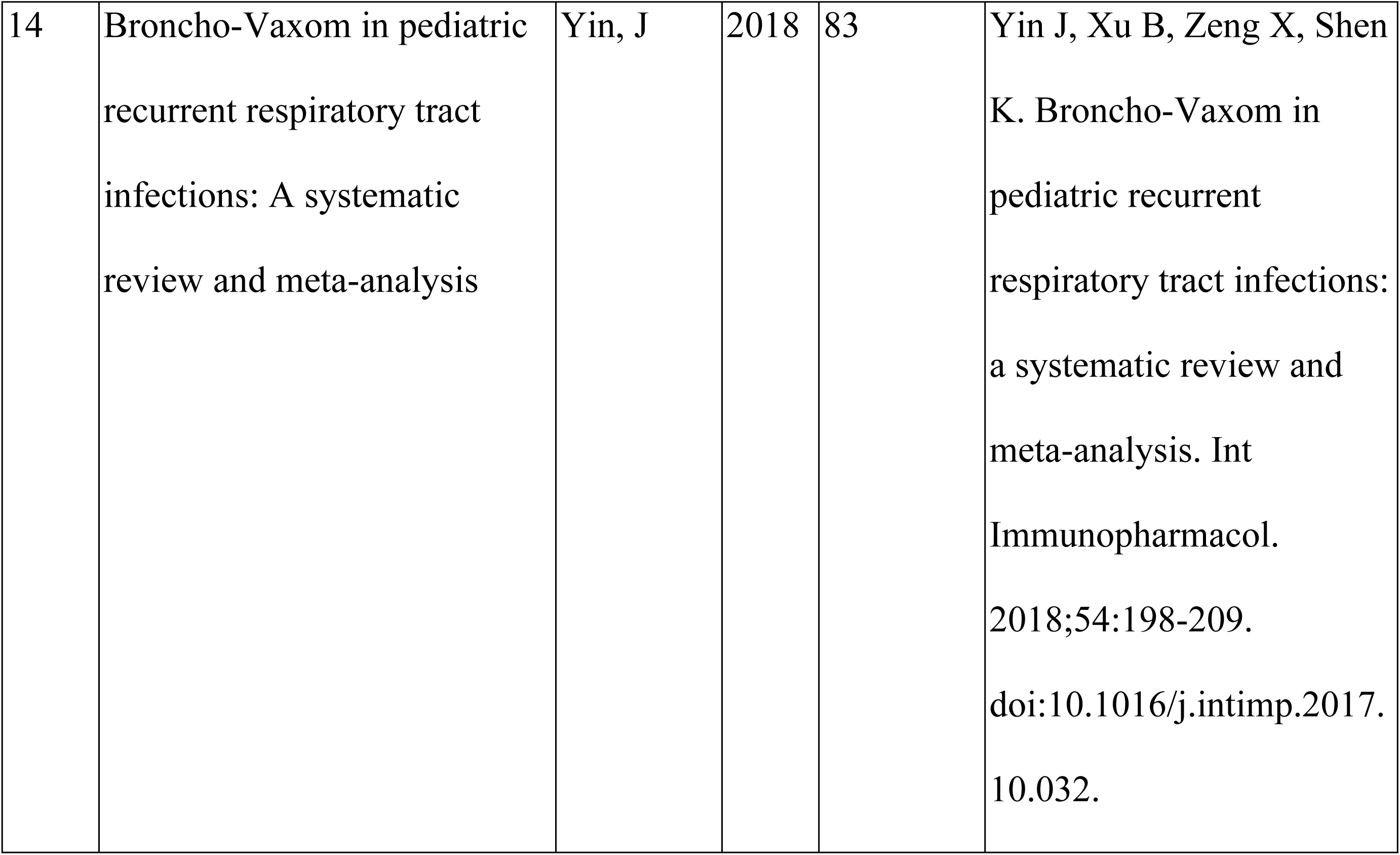
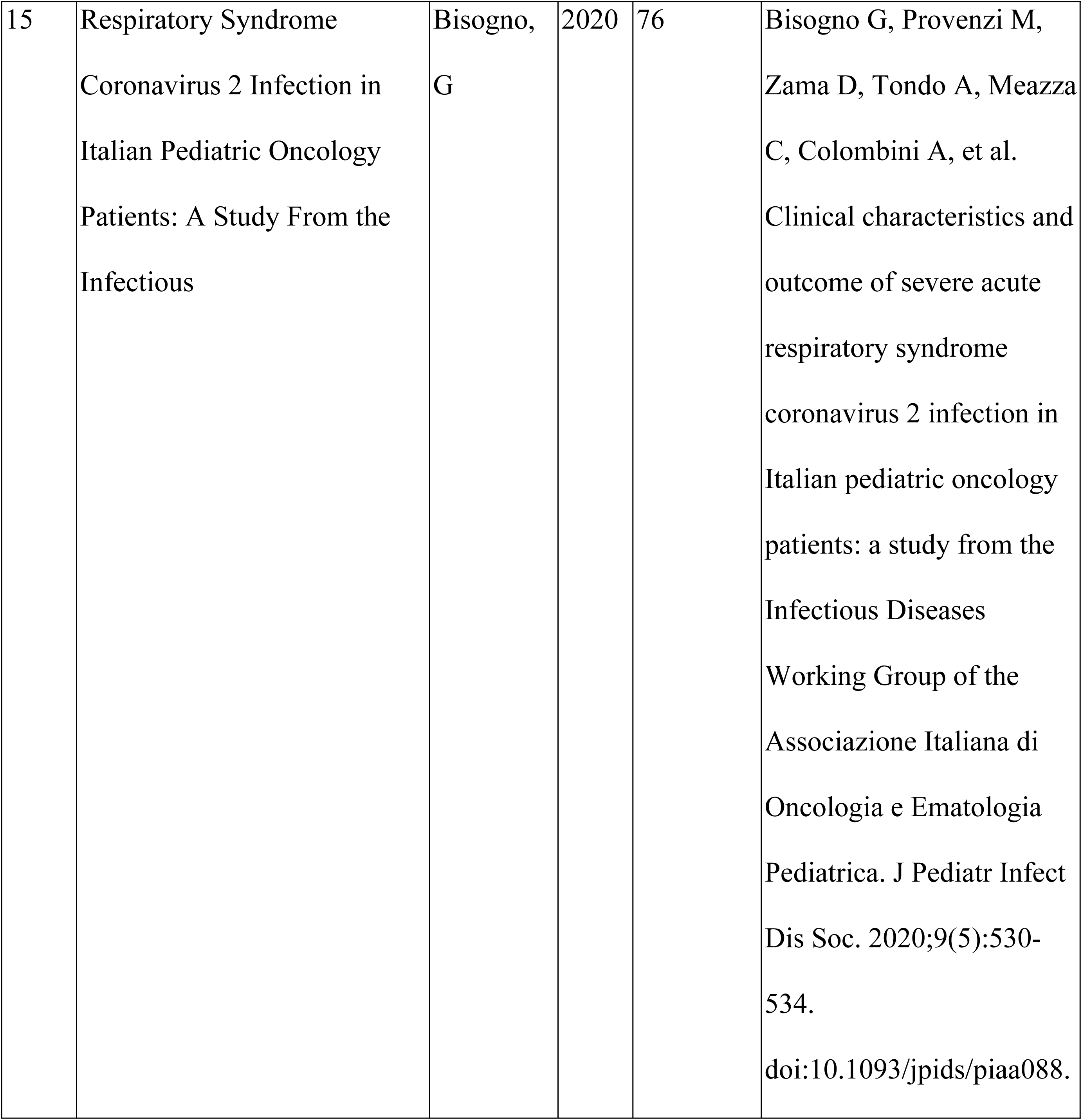
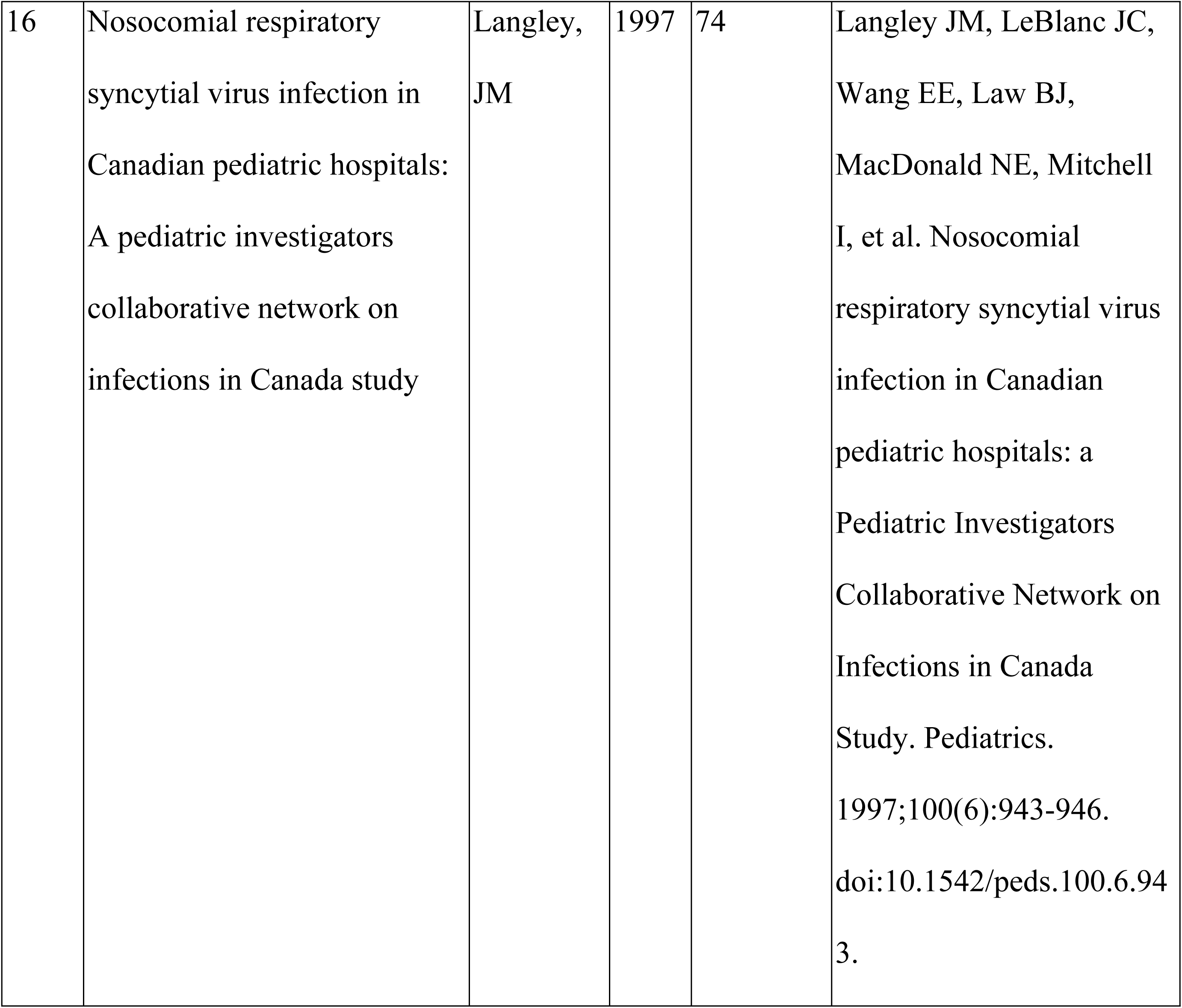
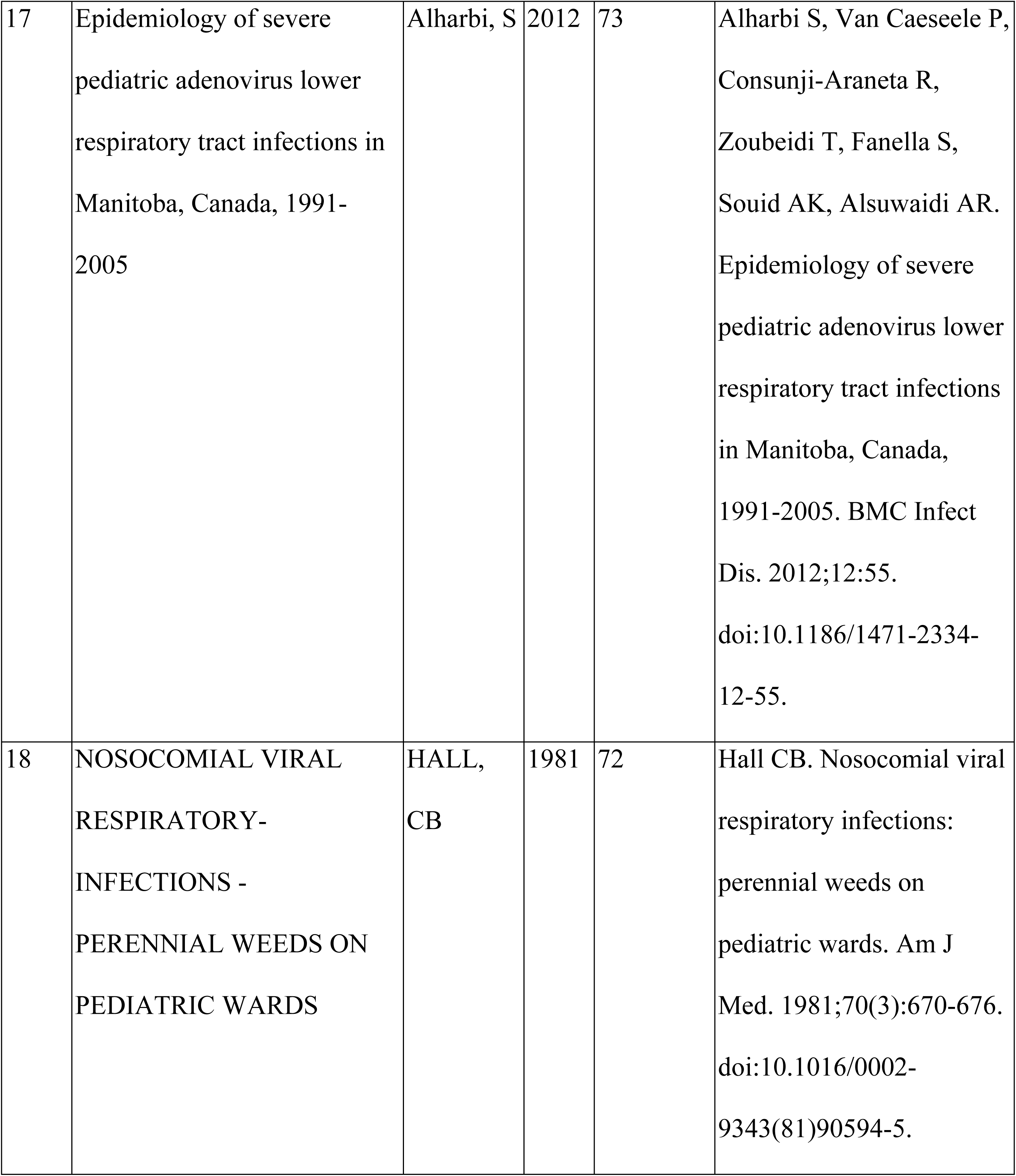
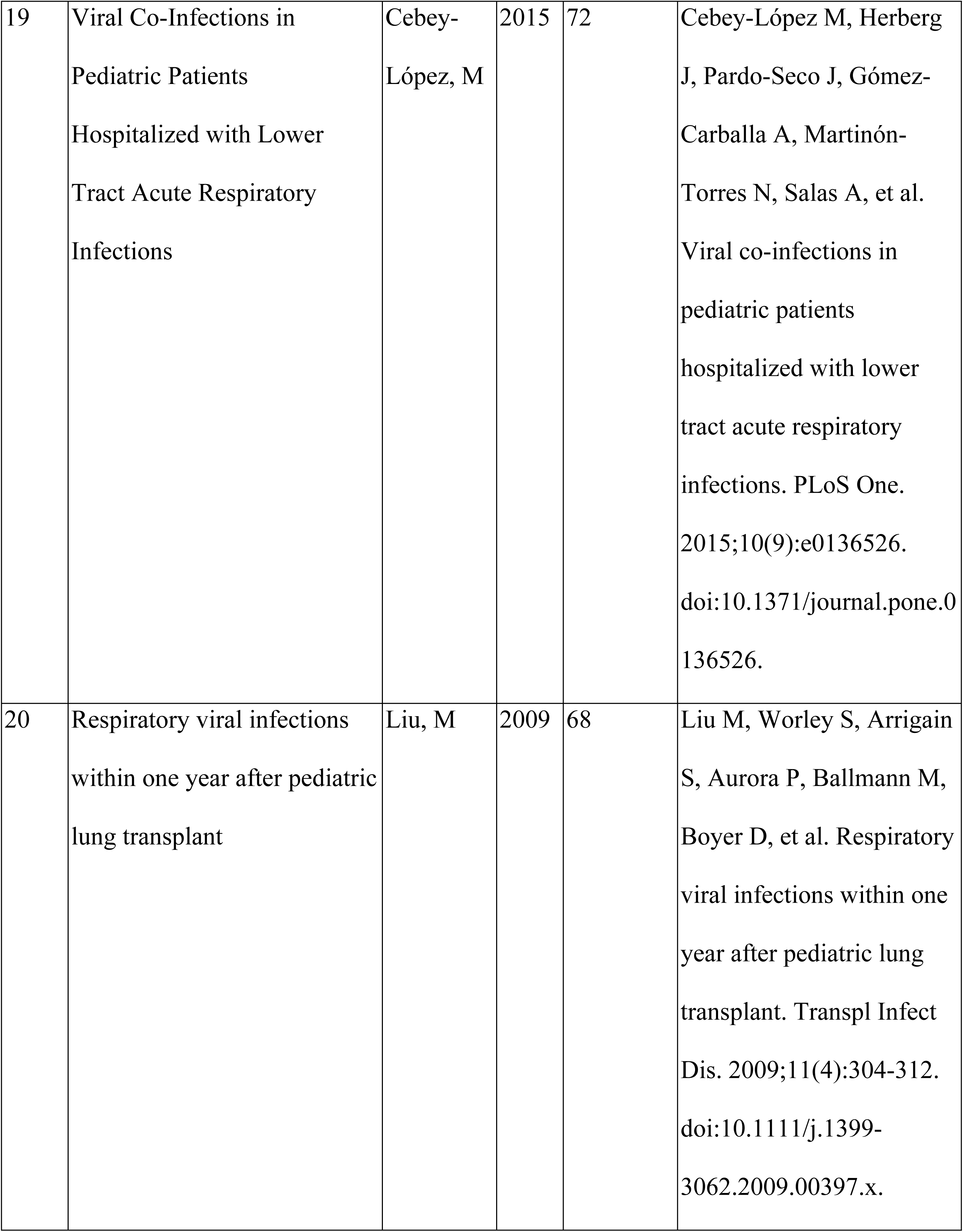
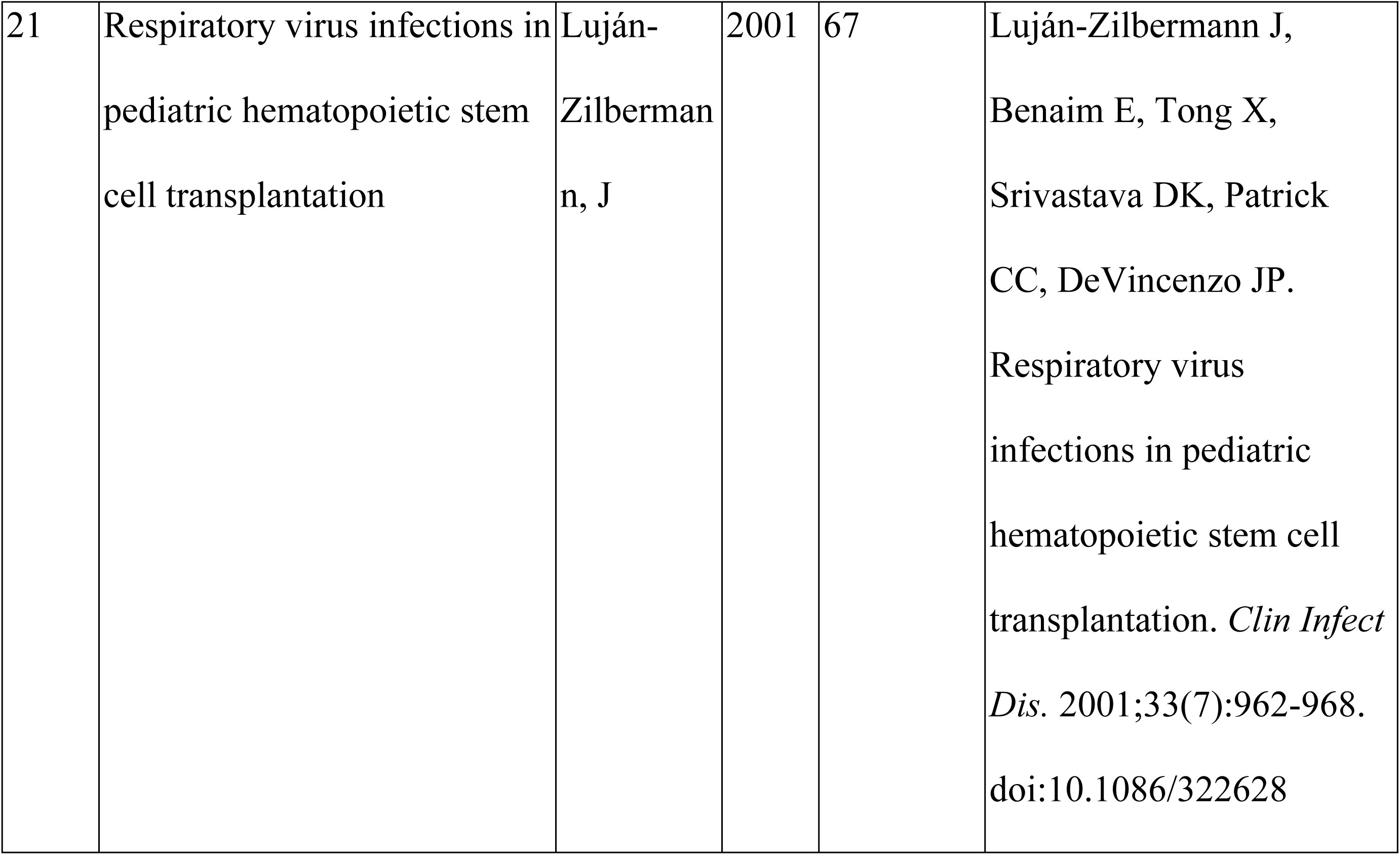
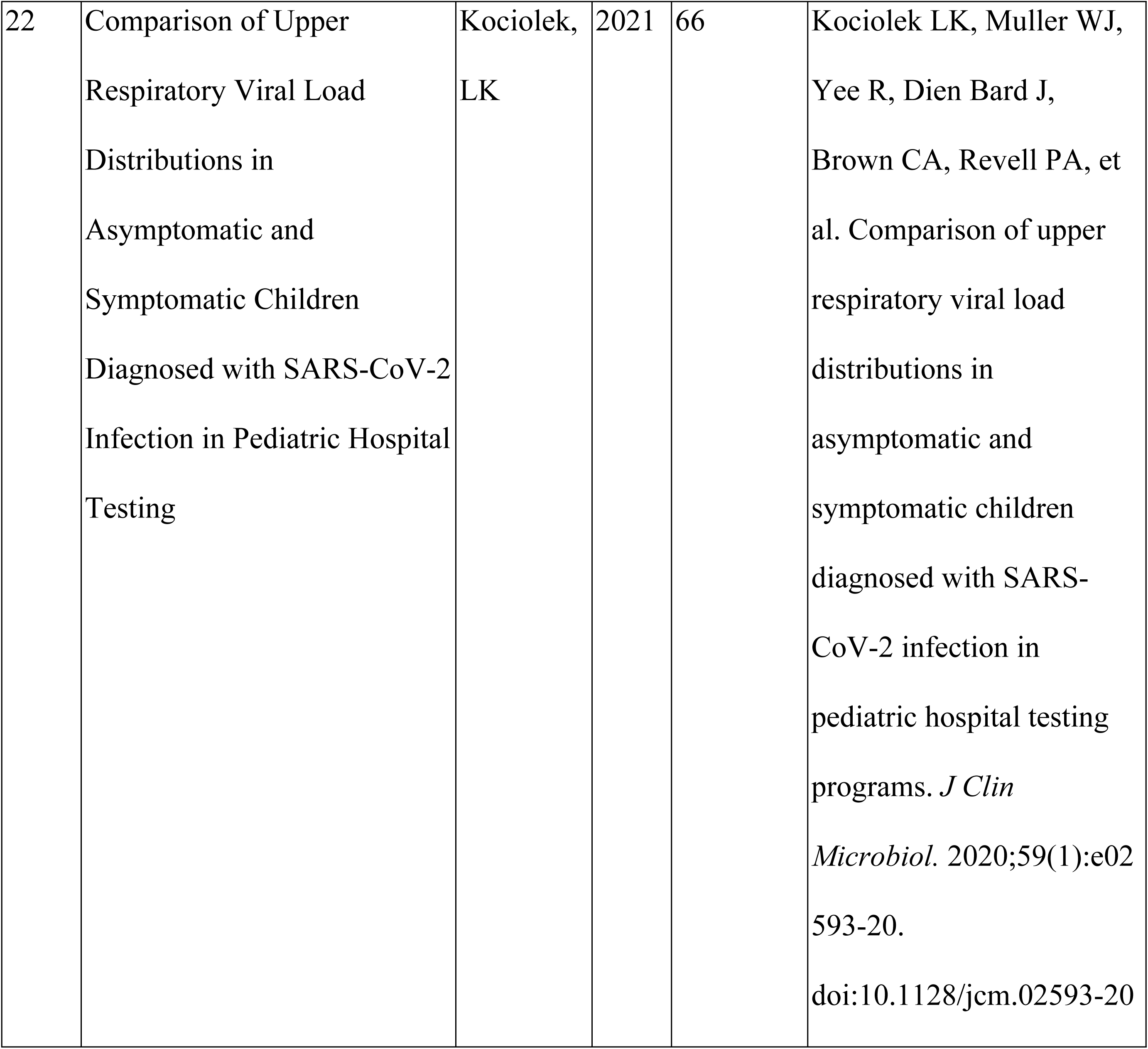
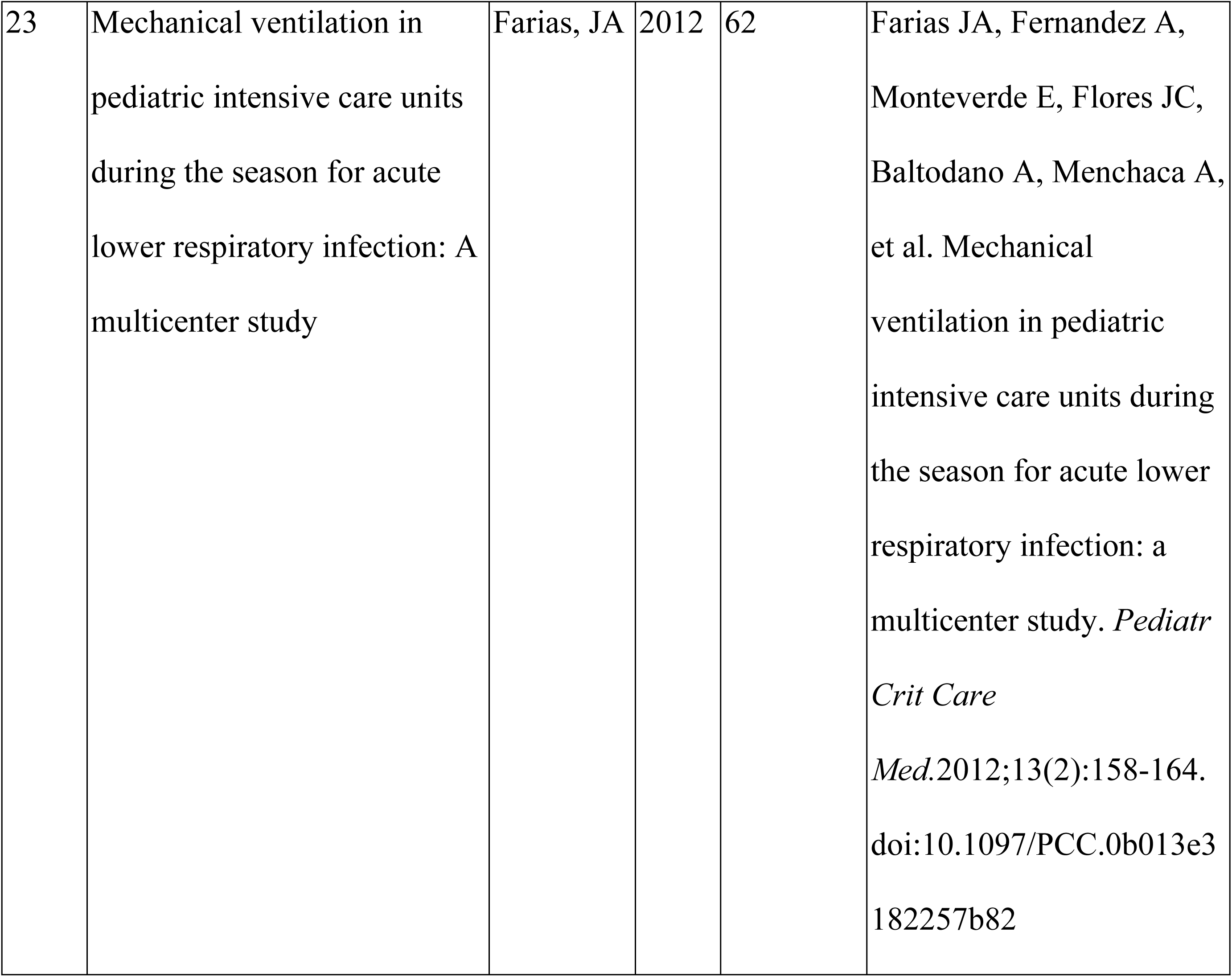
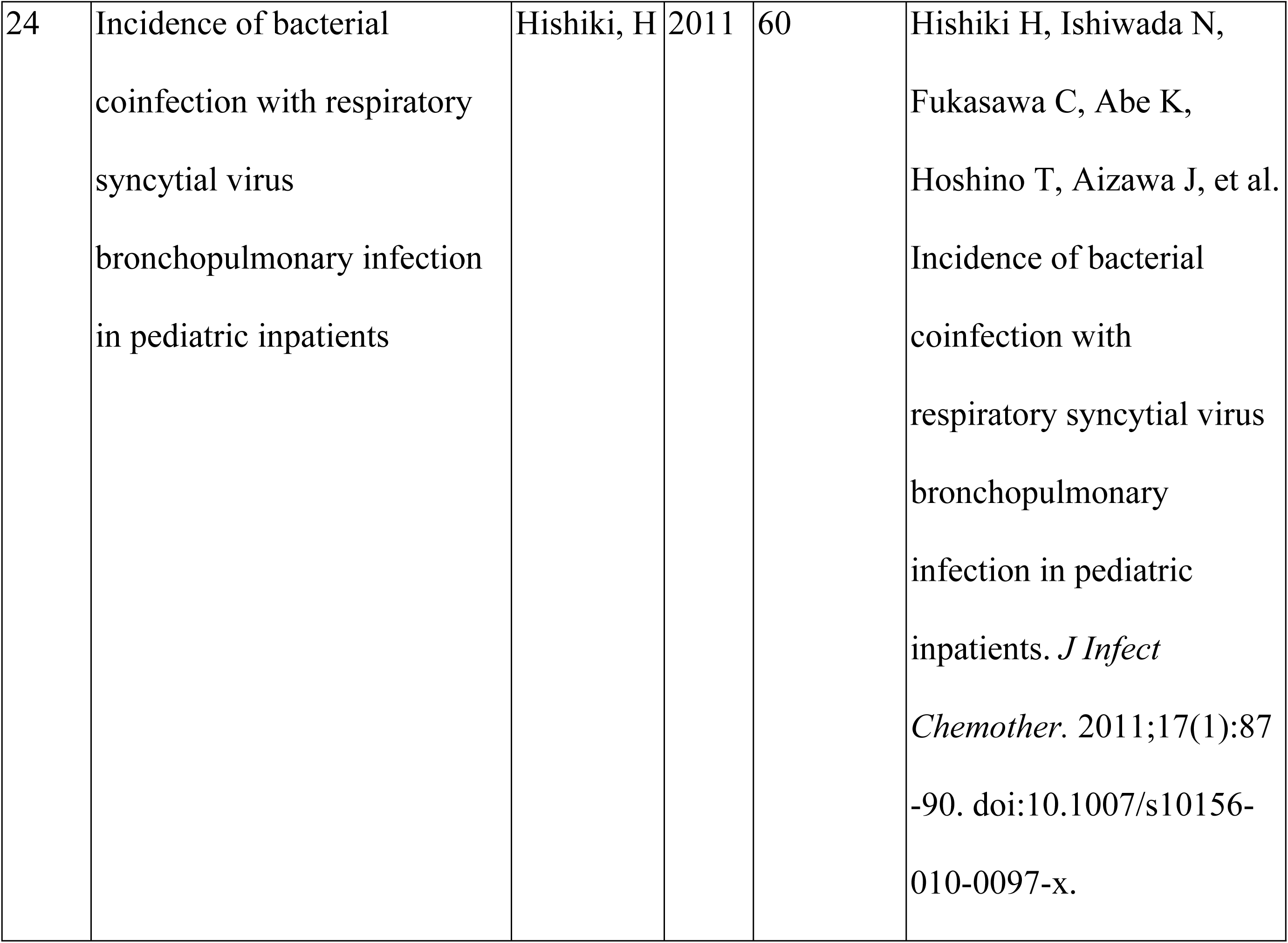
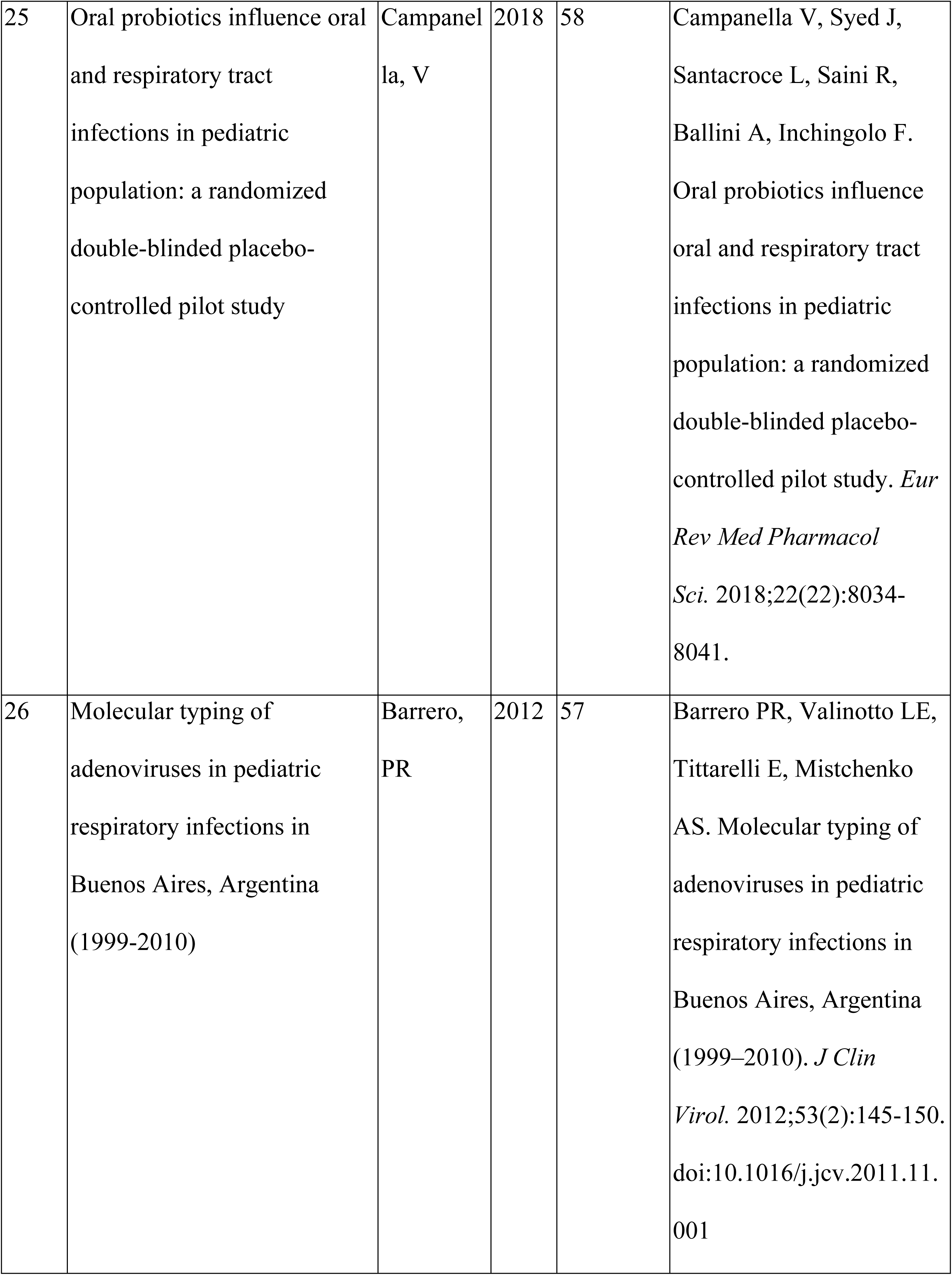
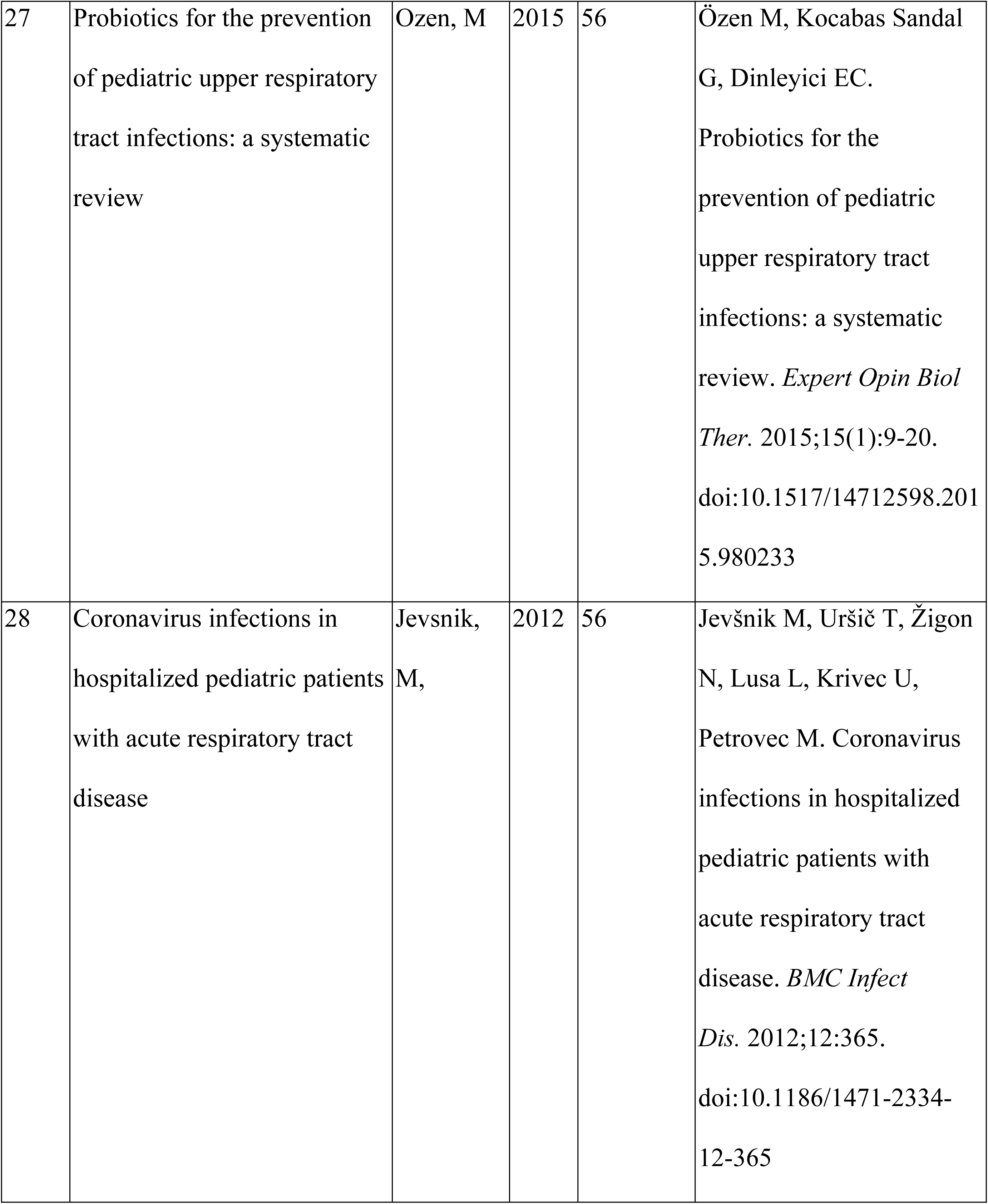
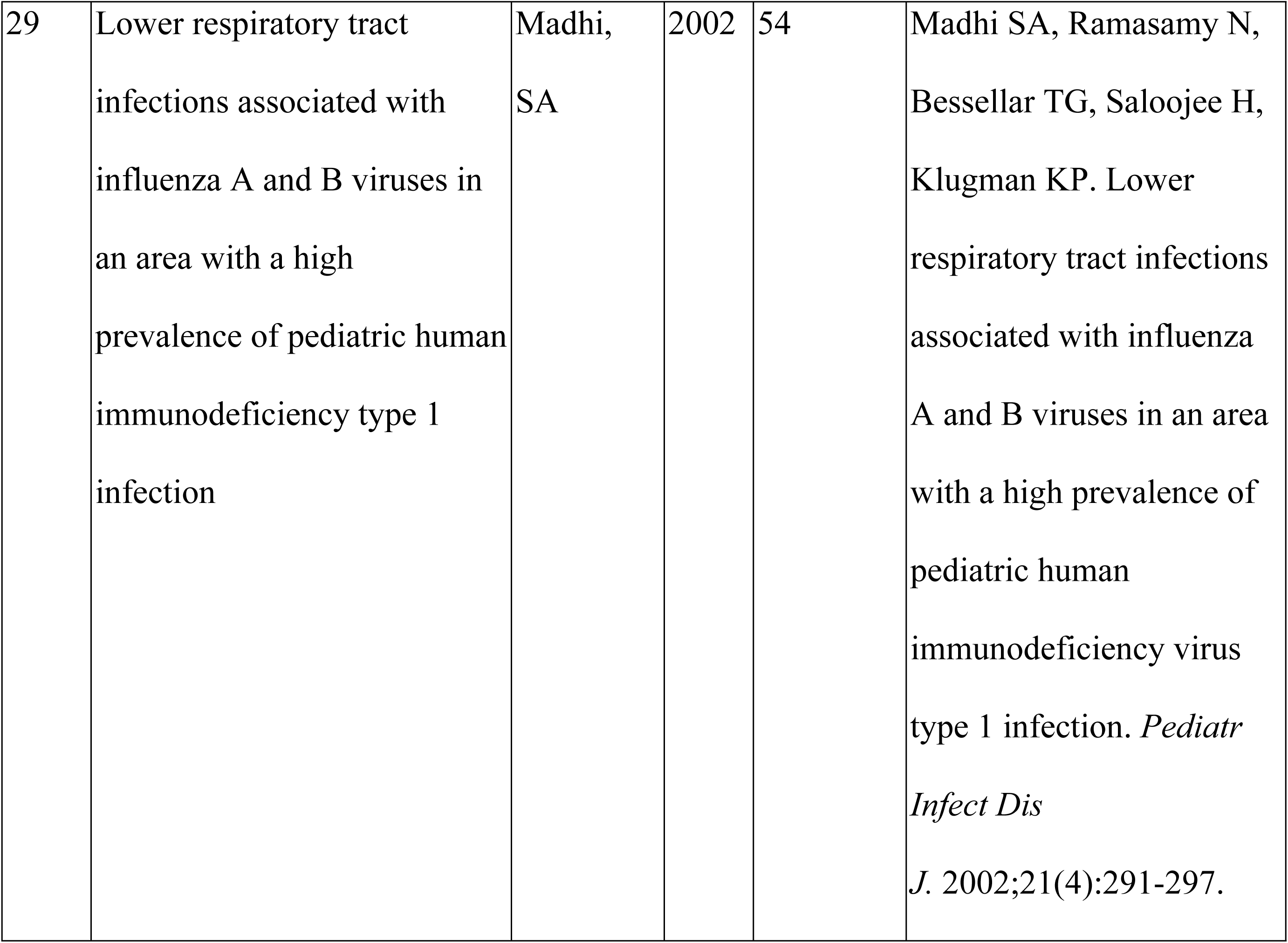
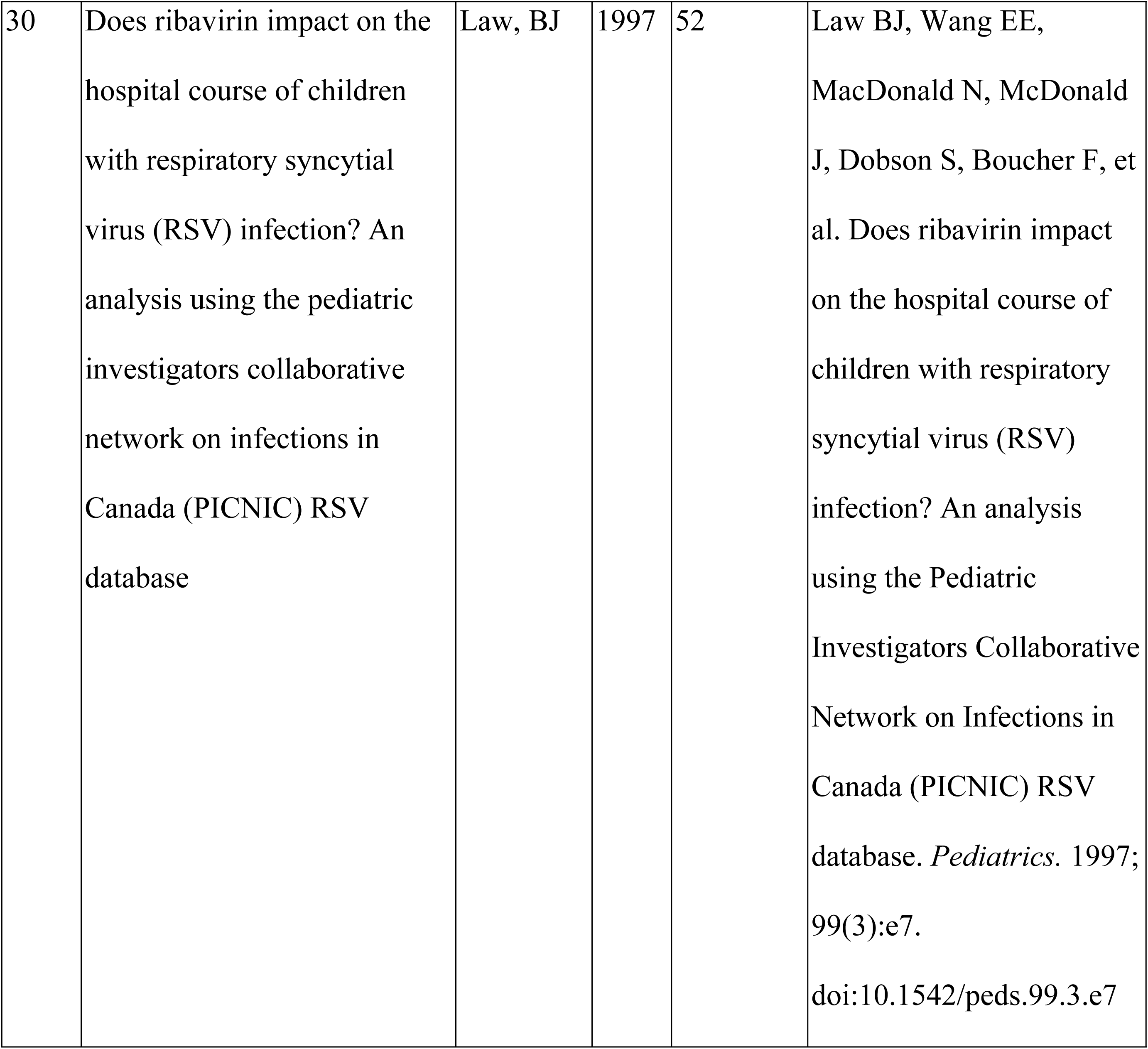
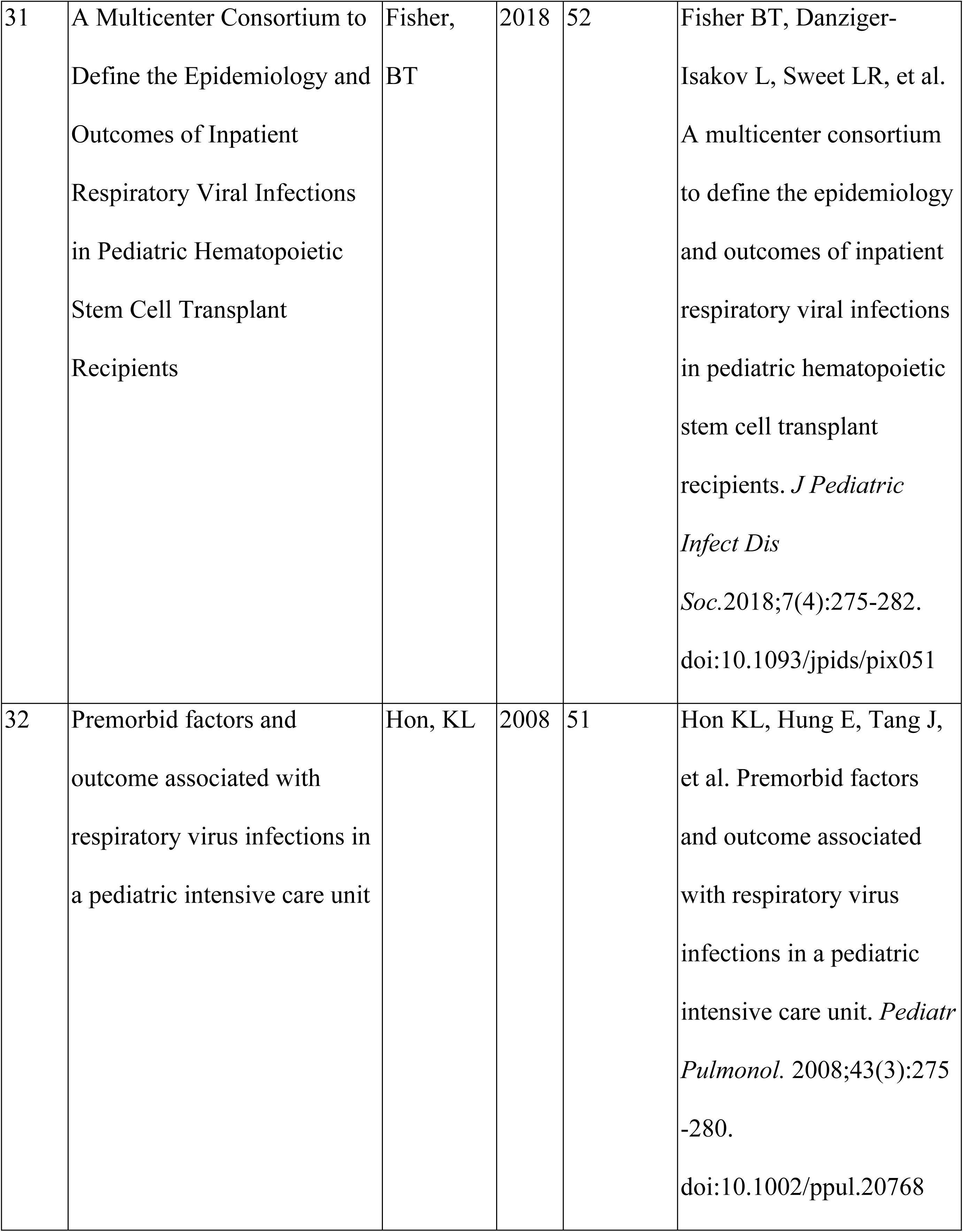
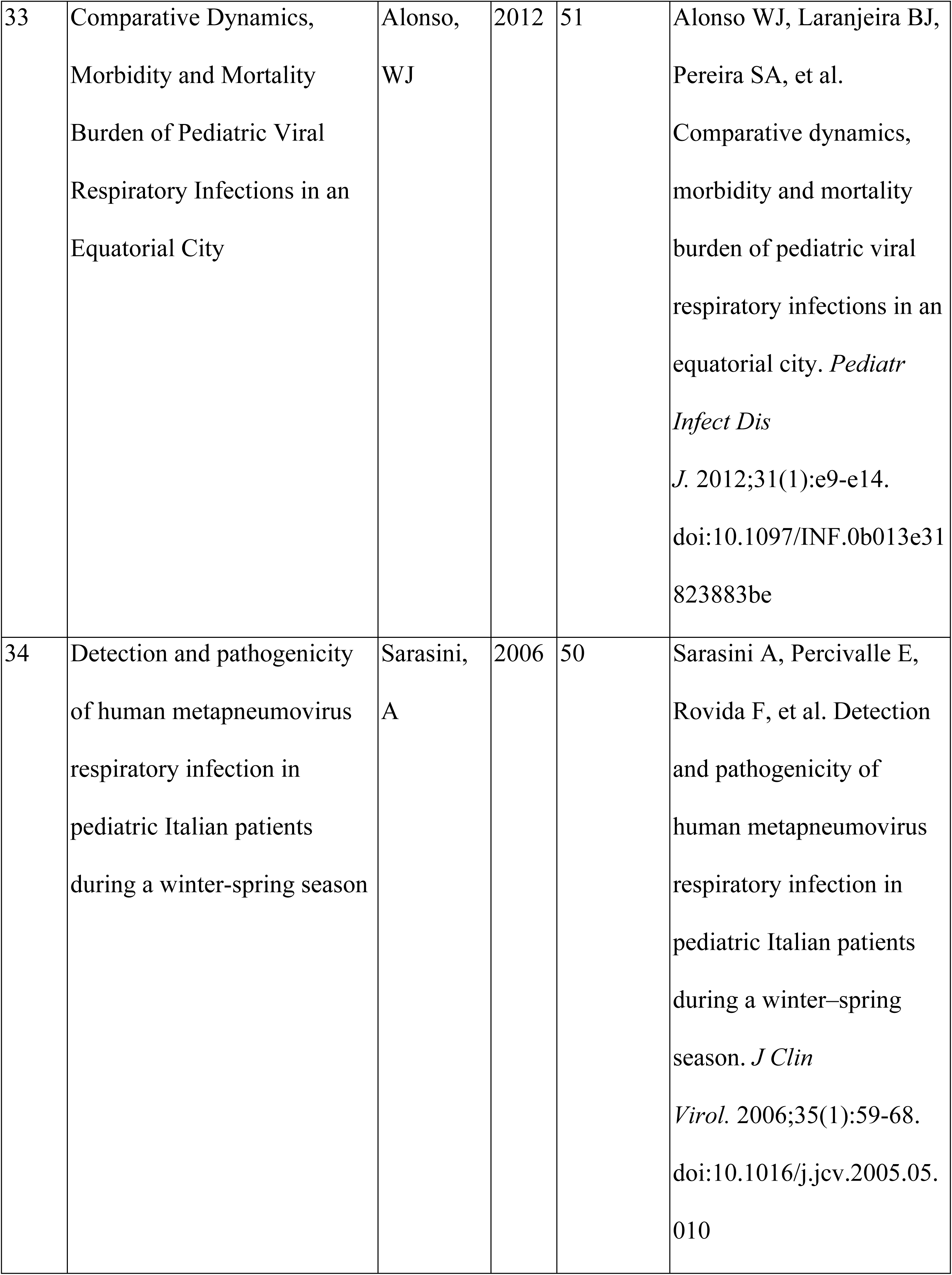
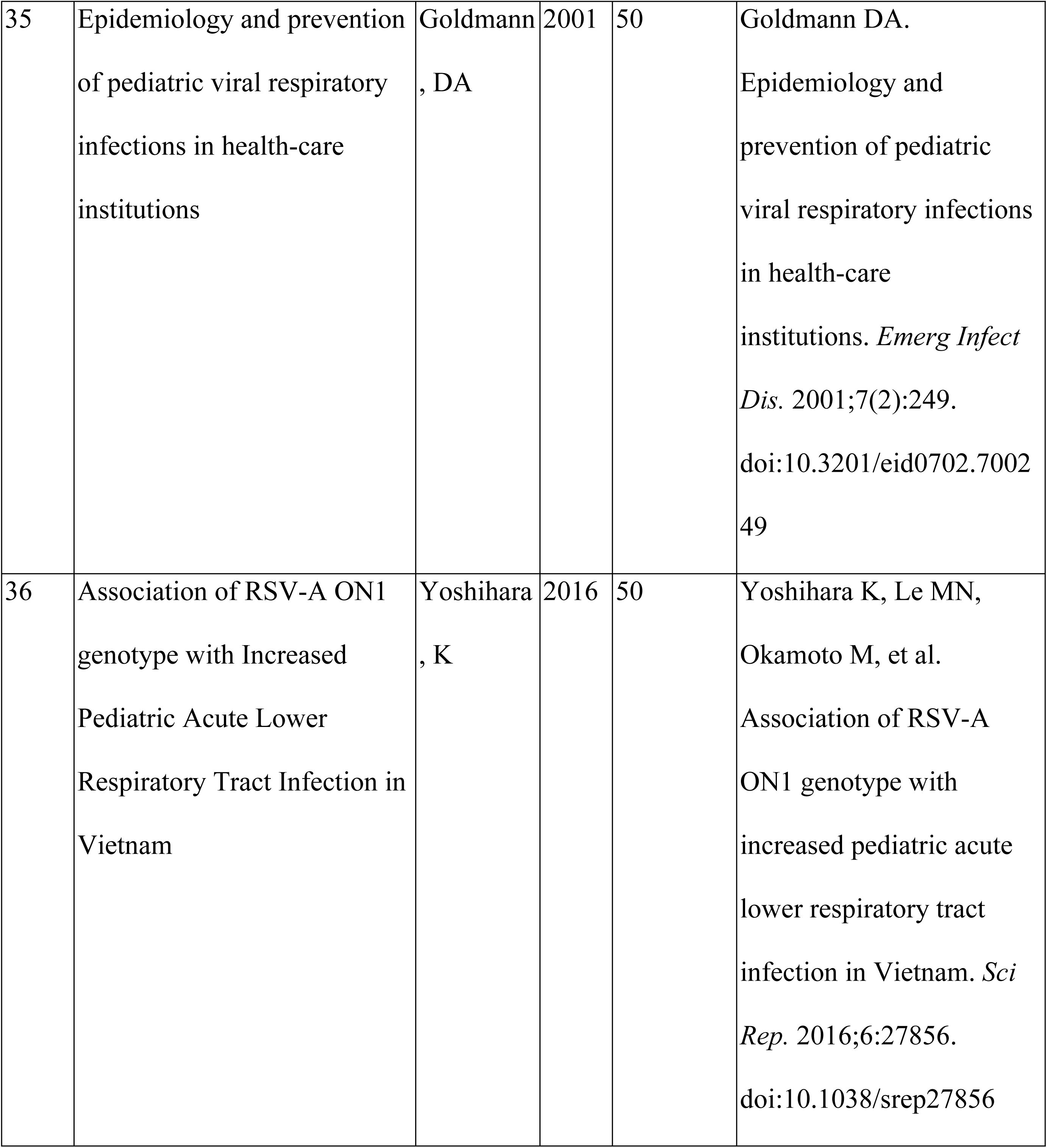
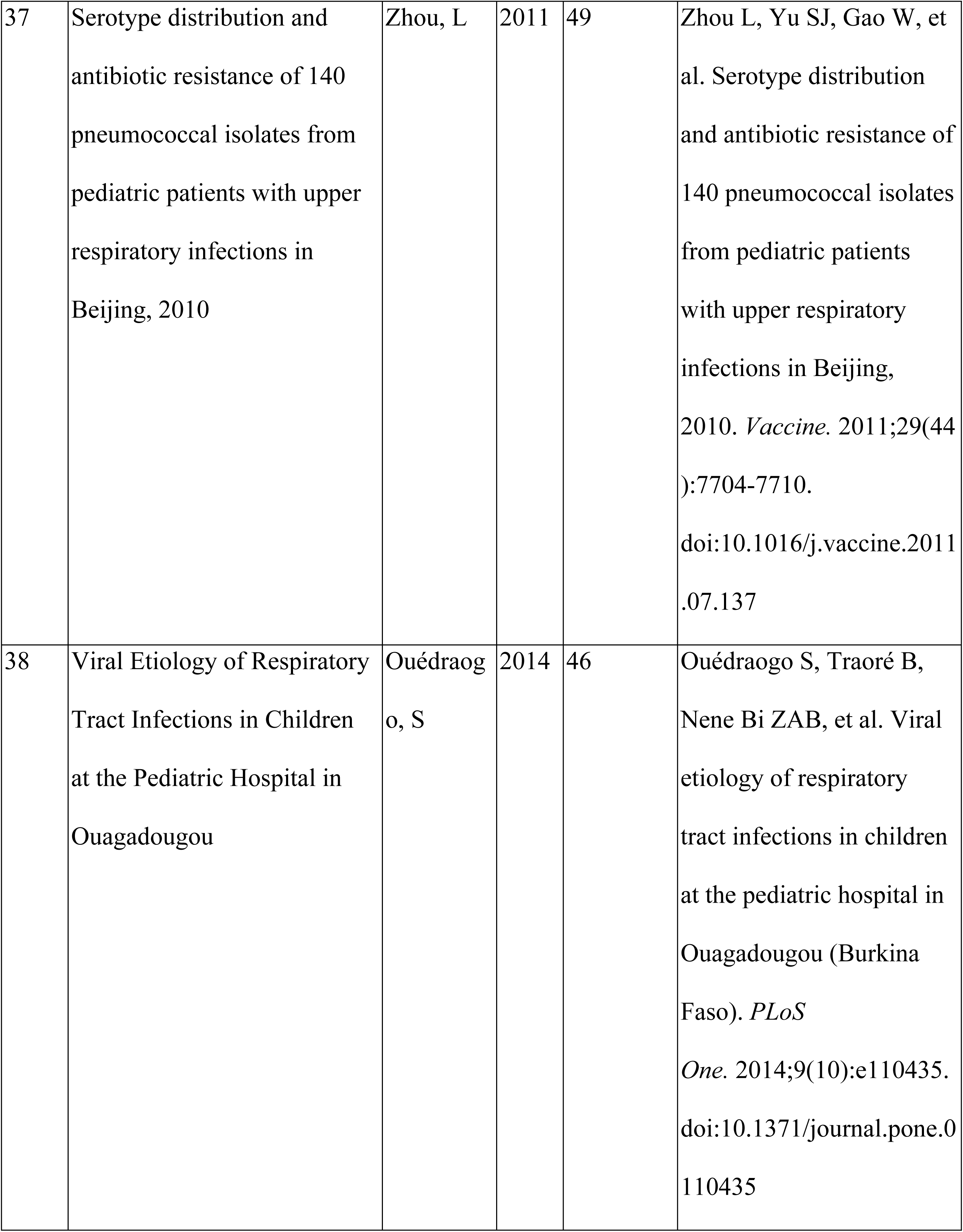
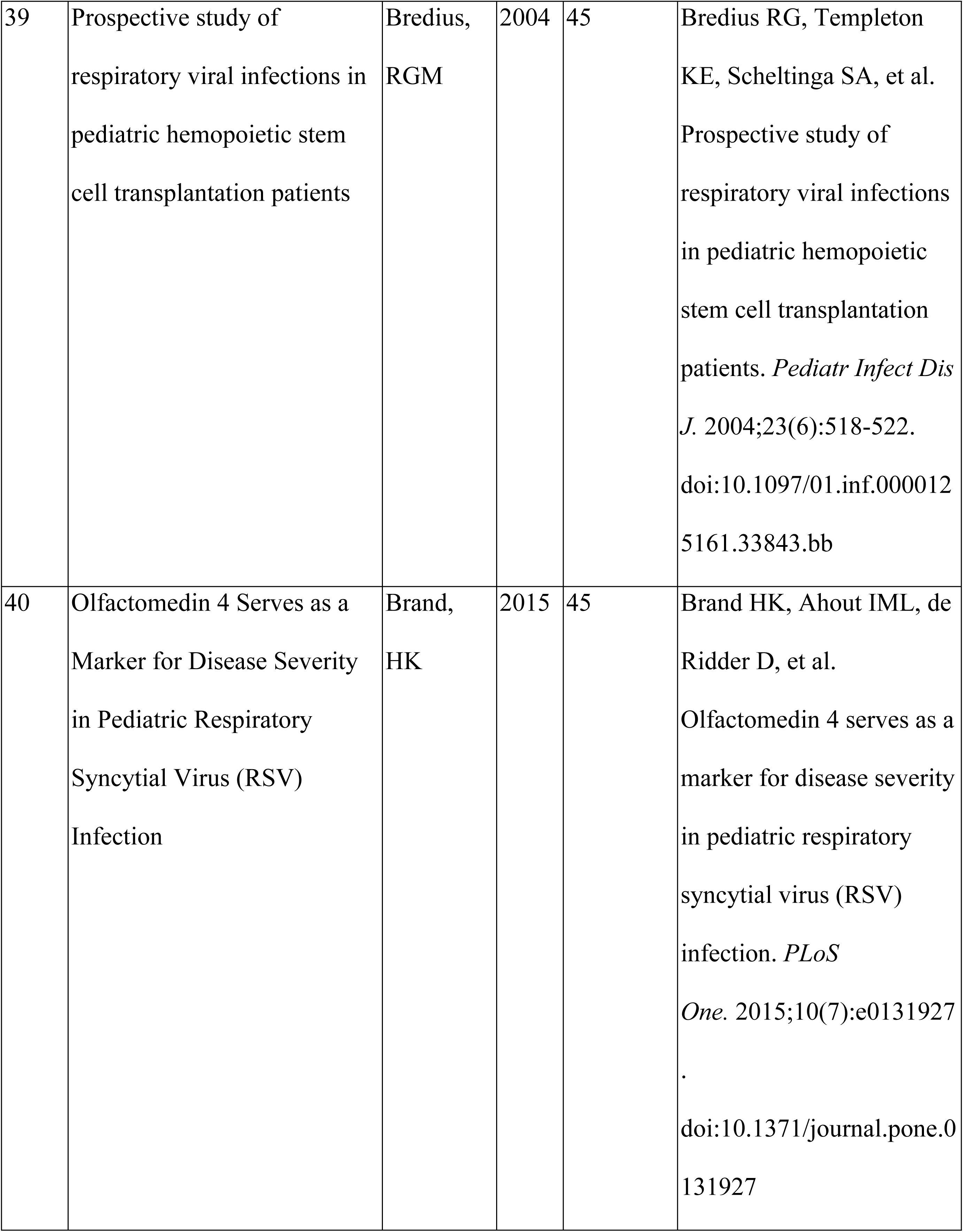
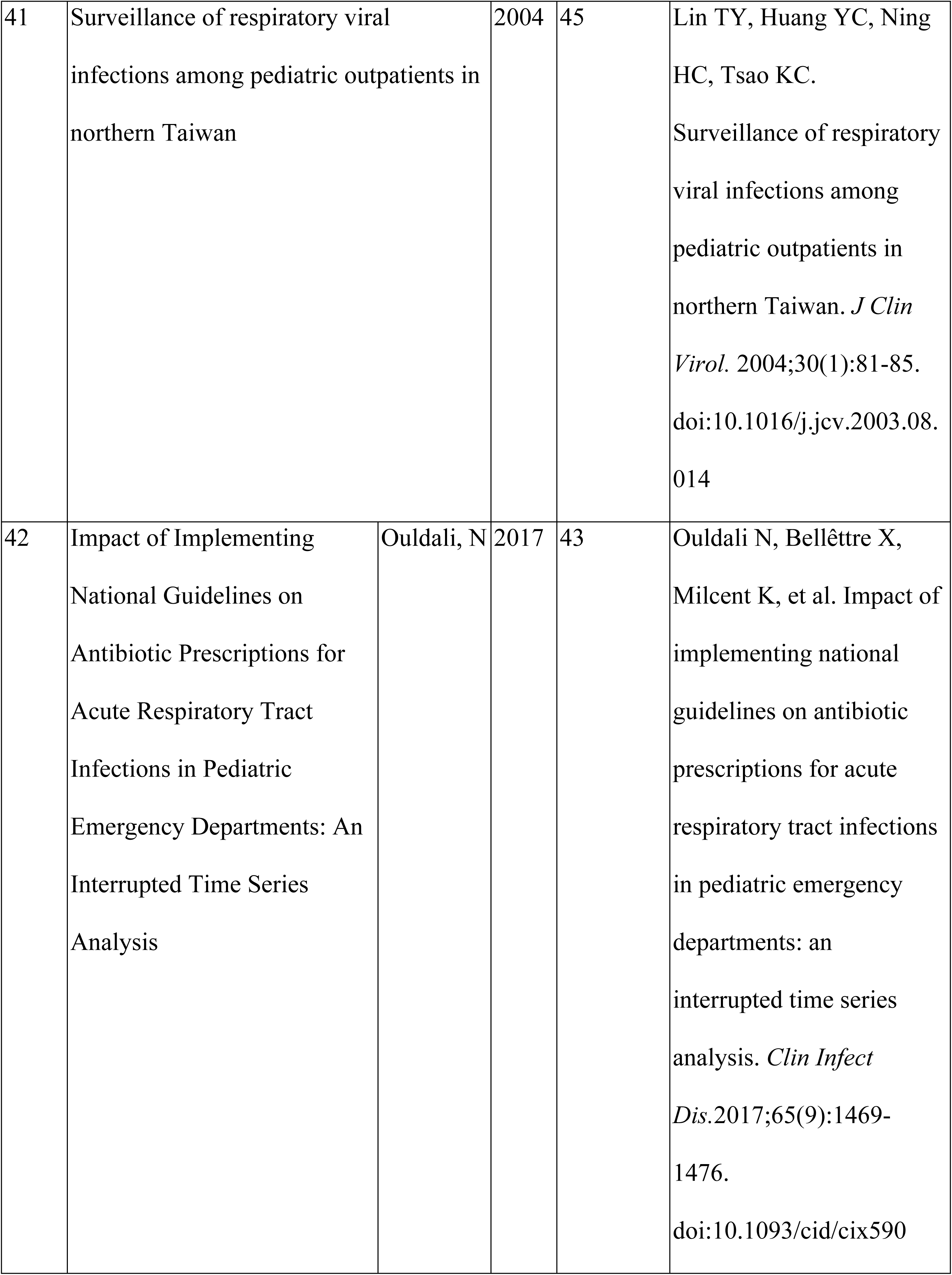
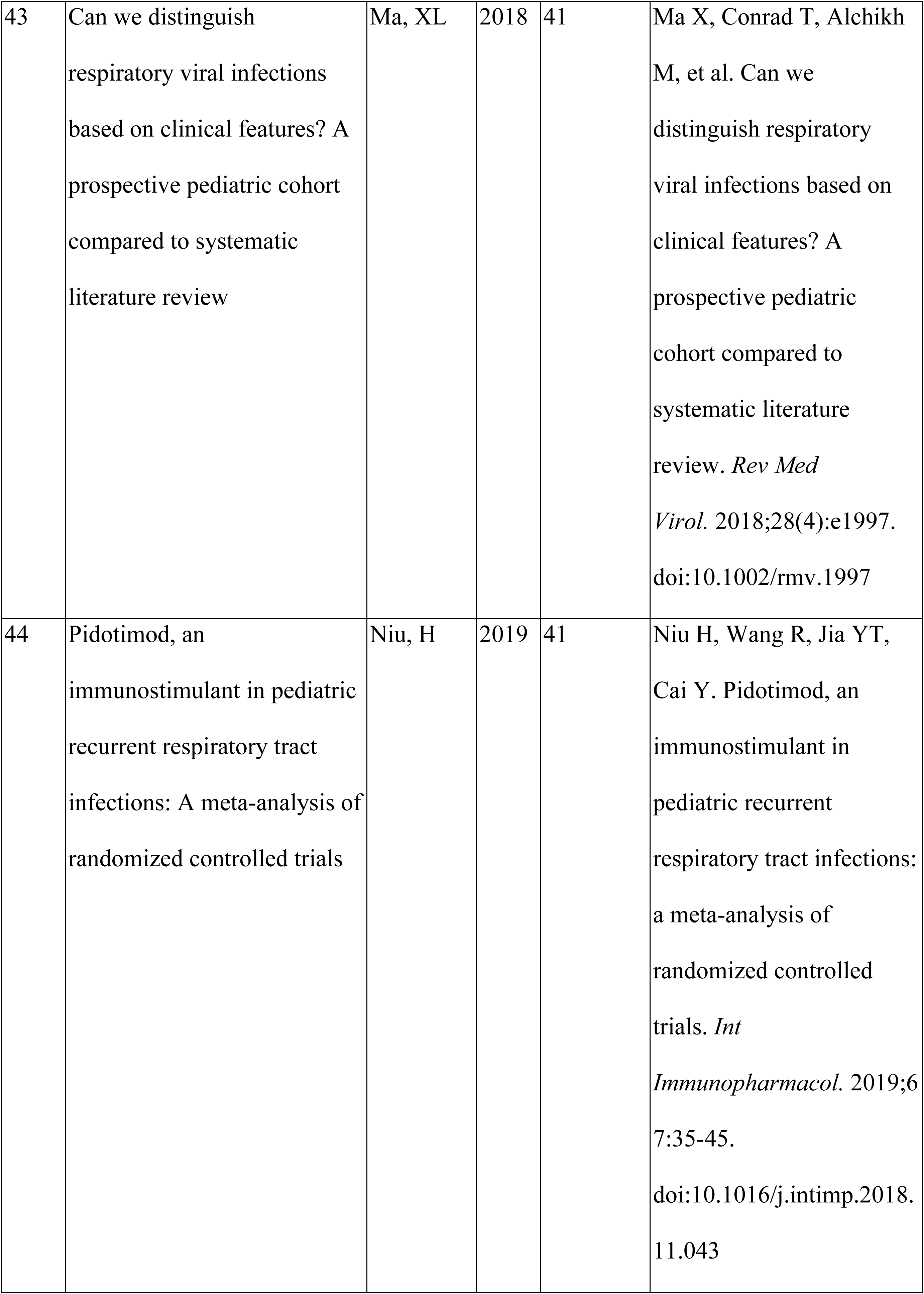
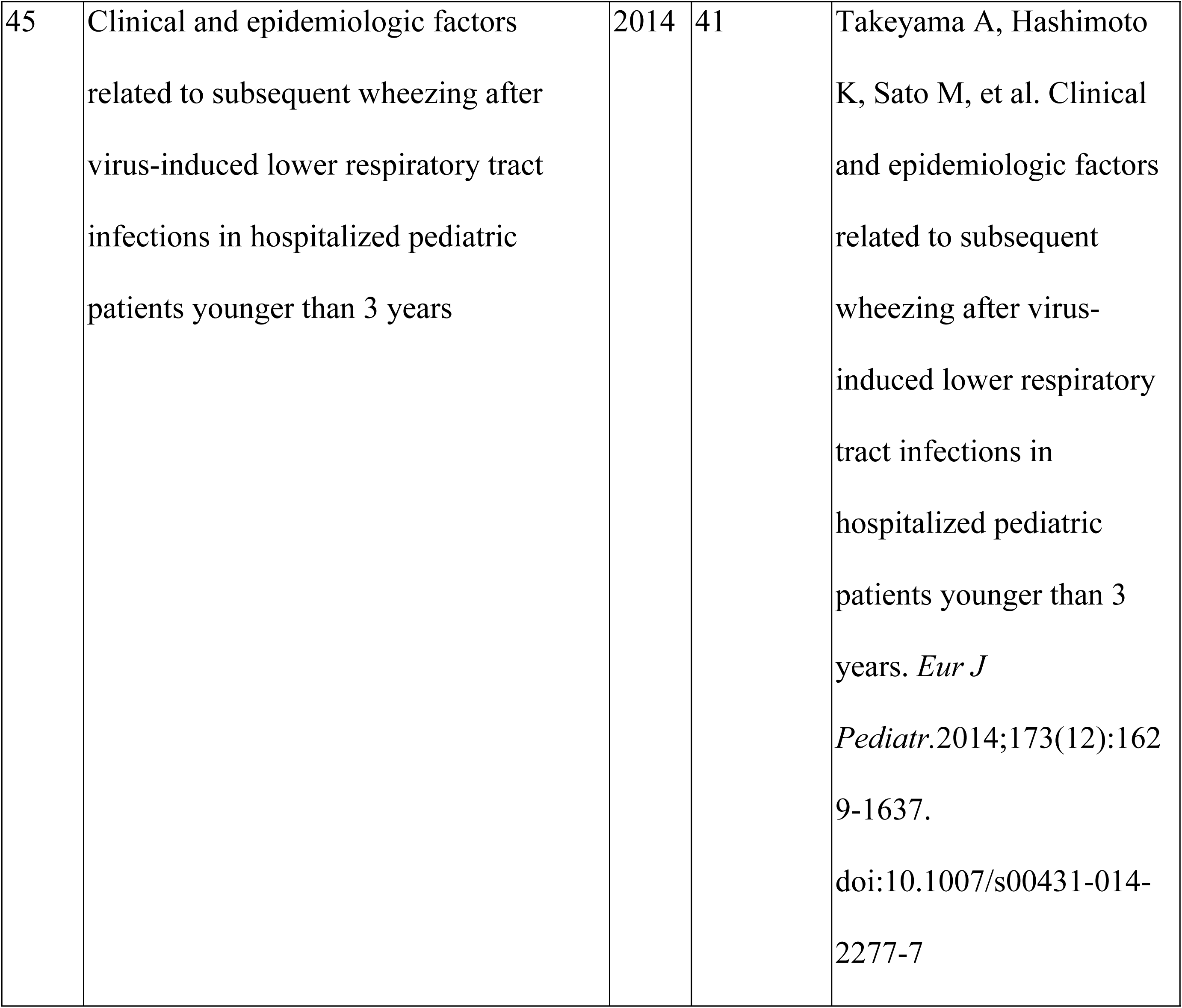
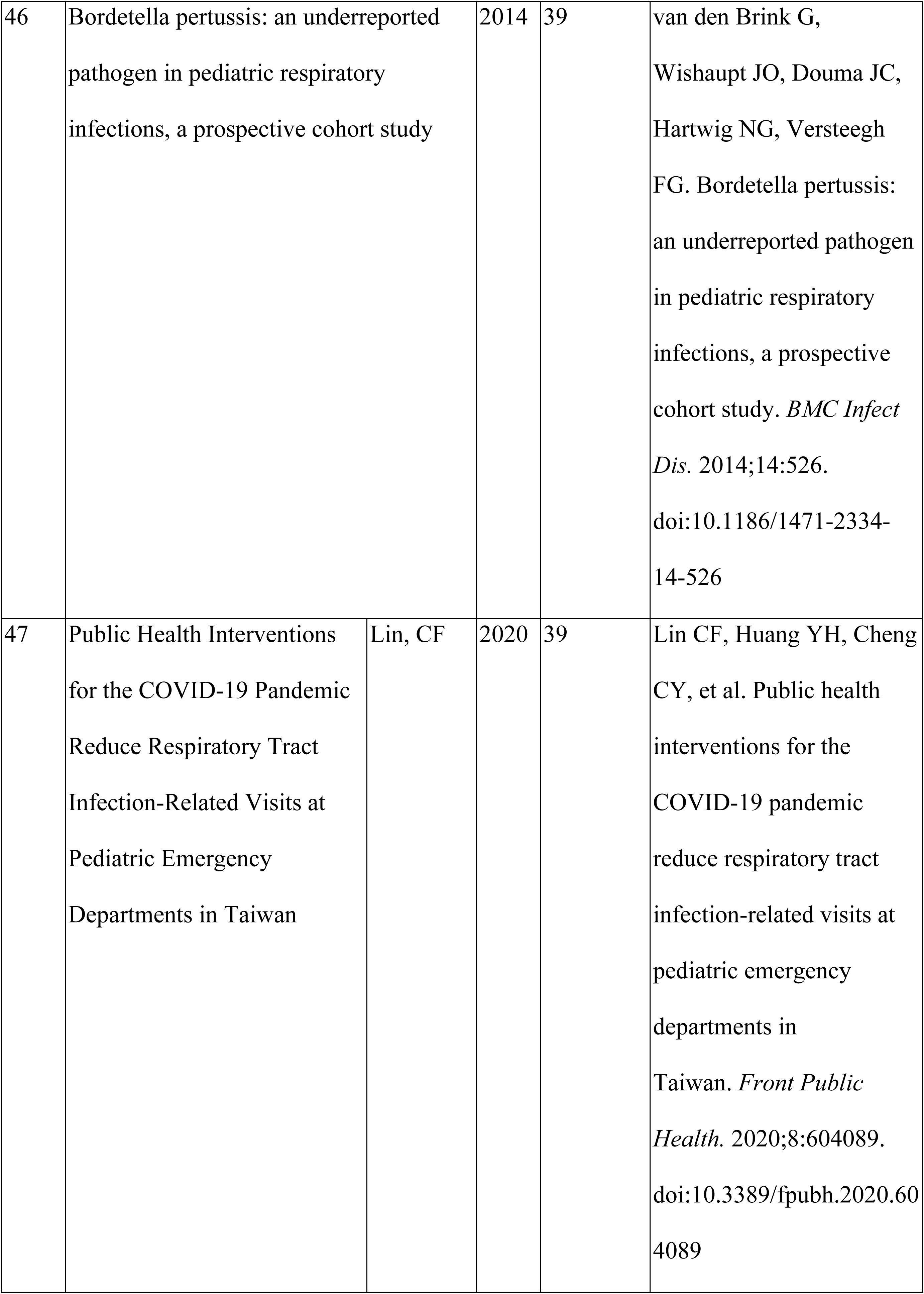
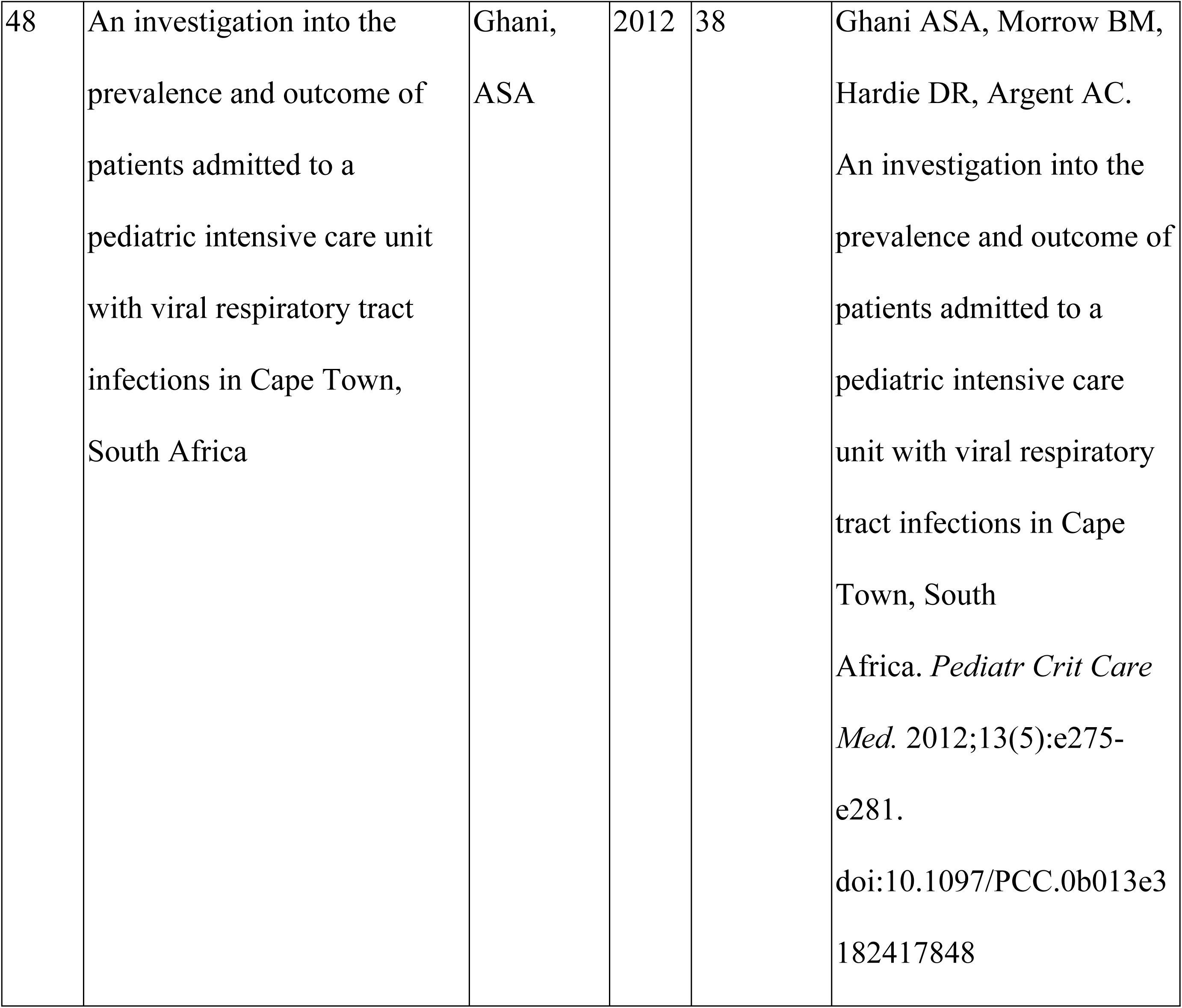
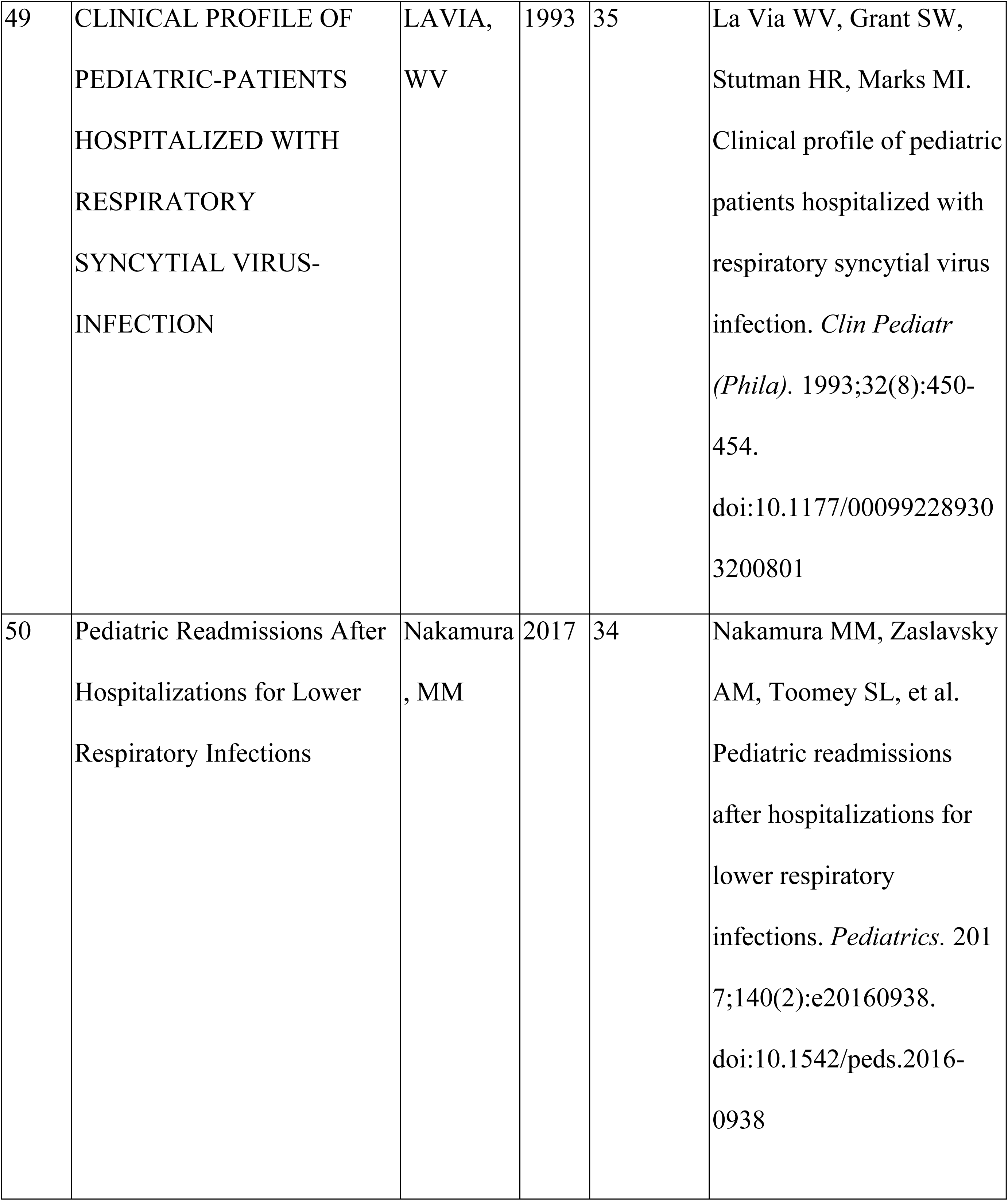
The 50 most-cited articles from Web of Science.

The temporal distribution, depicted in Table 2 and Figure 2, shows mid-to-recent concentration: 2010s account for 23/50 (46%), 2000s (12/50; 24%), and 1990s (9/50; 18%). Early seminal work is rare, with one study from the 1970s and one from the 1980s (1/50; 2%, each), while the 2020s represent 8% (4/50), consistent with the shorter time to accumulate citations. This suggests methodological development during the 1990s–2000s and peak citation visibility in the 2010s, with more recent contributions still accruing impact.

**Figure 2:**
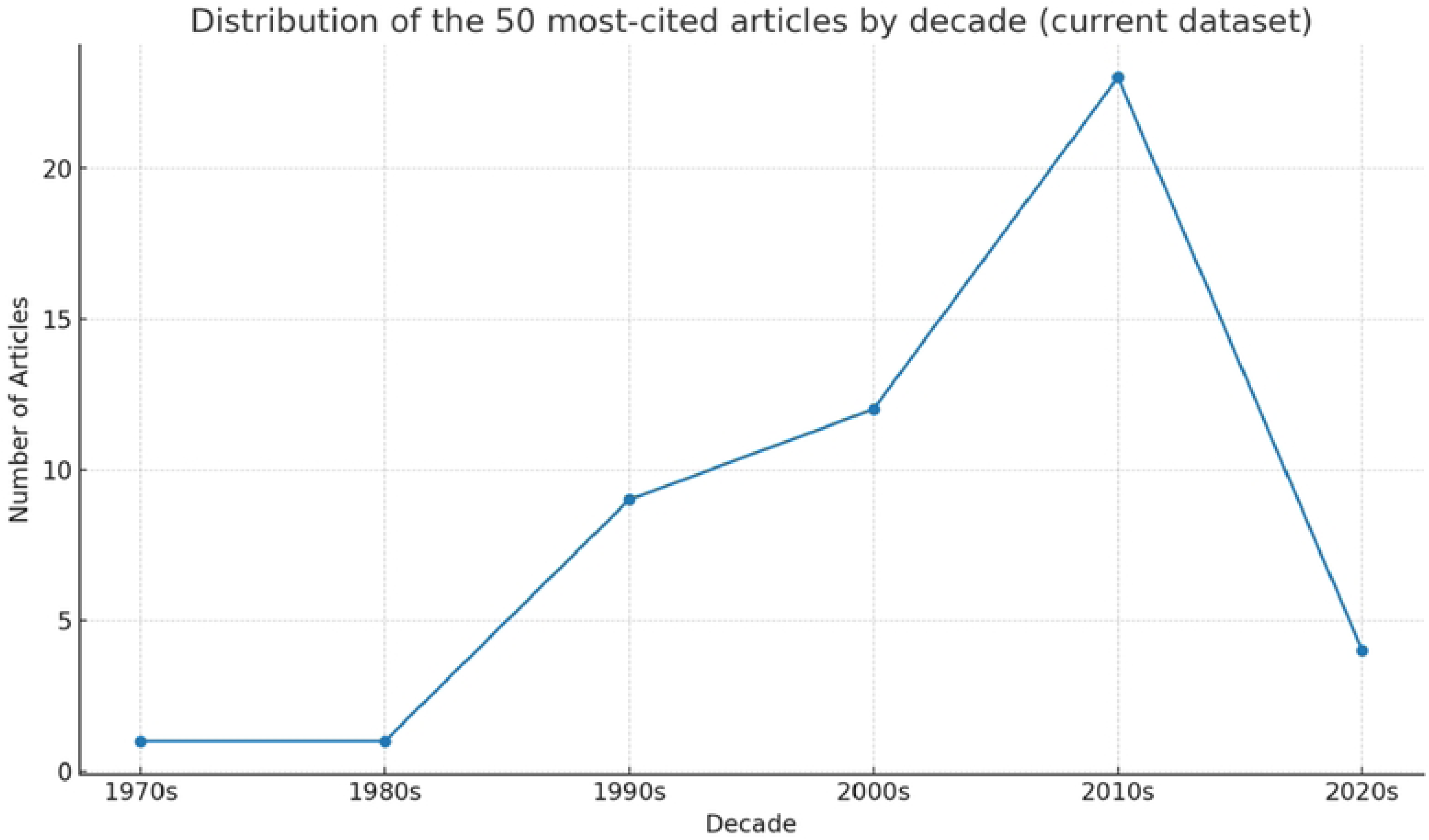
Article distribution by decade.

**Table 2:**
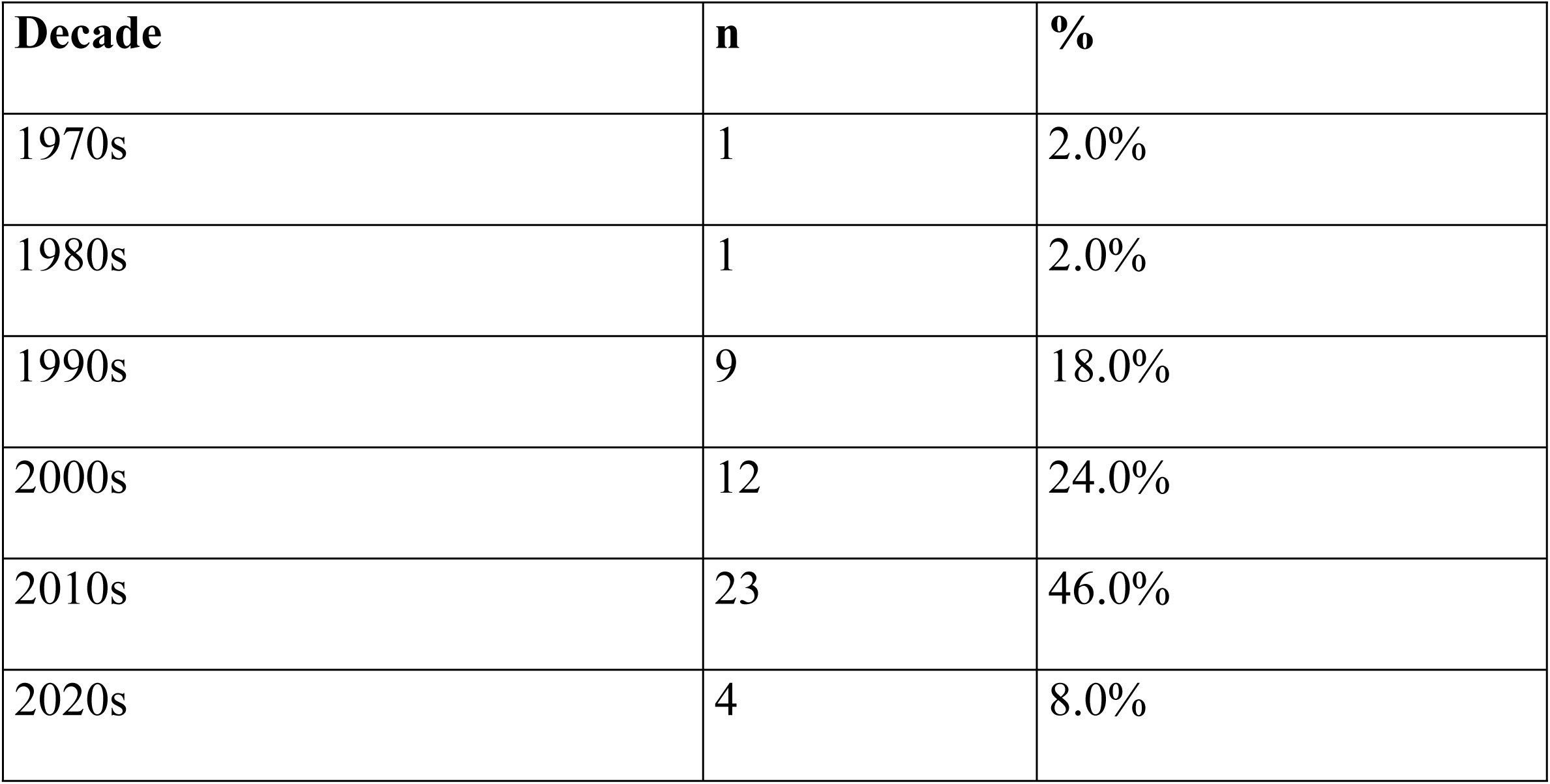
Article distribution by decade.

Figure 3 and Table 3 illustrate that study designs are dominated by observational cohorts: 36% (18/50) prospective, 30% (15/50) retrospective. Randomized controlled trials (RCTs) represent 12% (6/50), while synthesis papers are fewer: systematic review plus meta-analysis (3/50; 6%), systematic review (1/50; 2%), review articles (4/50; 8%), and cross-sectional designs 6% (3/50). This demonstrates a reliance on cohort evidence, with RCTs and formal syntheses present but less prevalent.

**Figure 3:**
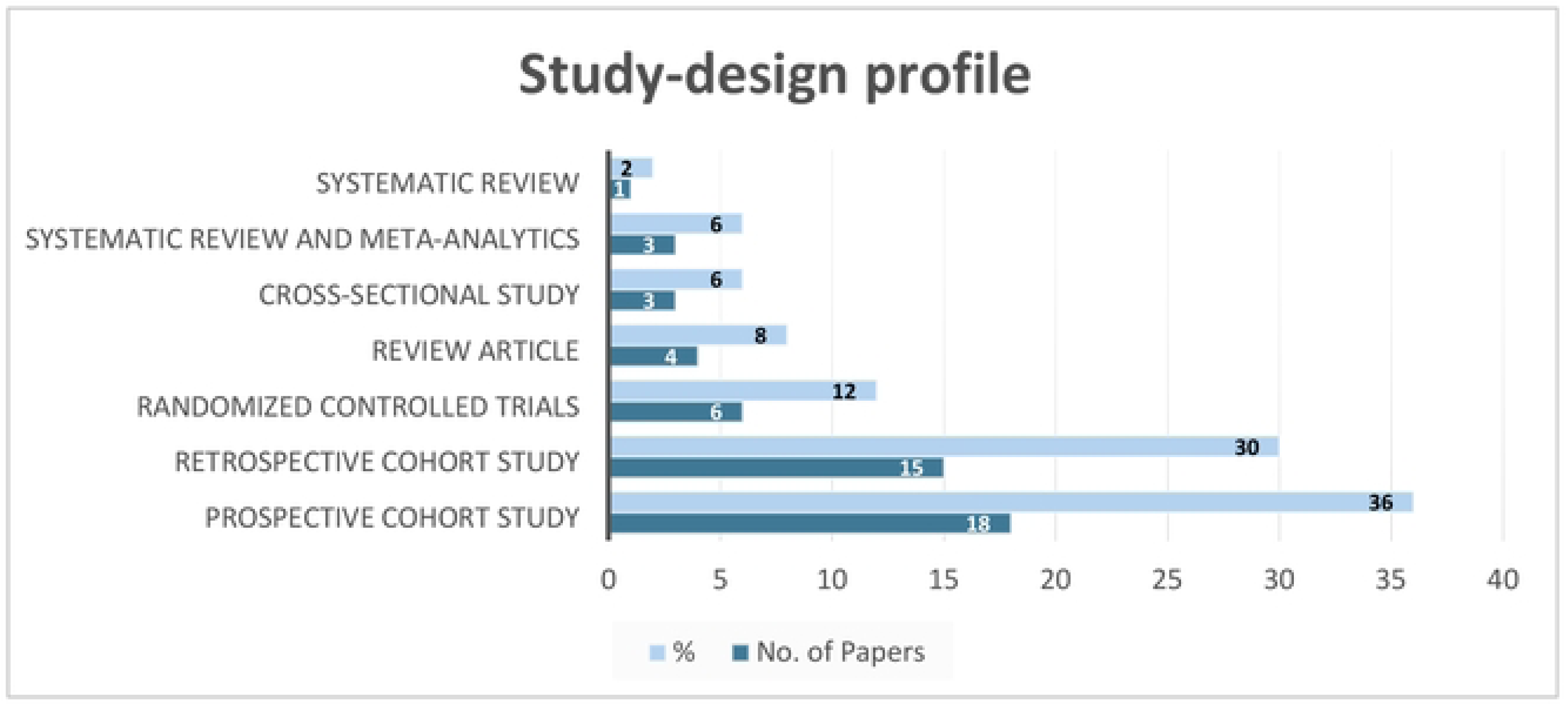
Study-design profile of the most-cited articles. Distribution of the 50 most-cited pediatric infection studies by study design, highlighting the predominance of observational cohort studies. Abbreviations: RCT = randomized controlled trial; Obs = observational study.

**Table 3:** Study-design profile of the most-cited articles.

| Study Design | No. of Papers | % |
| --- | --- | --- |
| Prospective Cohort Study | 18 | 36.0 |
| Retrospective Cohort Study | 15 | 30.0 |
| Randomized Controlled Trials | 6 | 12.0 |
| Review Article | 4 | 8.0 |
| Cross-Sectional Study | 3 | 6.0 |
| Systematic Review and Meta-Analytics | 3 | 6.0 |
| Systematic Review | 1 | 2.0 |

Publication across journals is somewhat dispersed, with the 50 papers in 31 journals. Six journals dominate: Pediatrics (n = 5; 10%), Pediatric Infectious Disease Journal (4; 8%), BMC Infectious Diseases and Journal of Clinical Virology (3 each; 6%), and Journal of Pediatrics and Journal of Clinical Microbiology (2 each; 4%). These six journals account for 38% of publication, with the remaining 25 journals contributing 62%. Although Pediatrics leads by article count, Journal of Pediatrics commands the largest cumulative citation count per paper, illustrating how highly visible journals concentrate impact (Table 4).

**Table 4:** Top journals contributing to the 50 most-cited articles (current dataset). * JCR 2024 reflects the **2023 Journal Impact Factor** values released in the 2024 Journal Citation Reports.

| **Rank** | **Journal** | **No. of Papers** | **2024 JCR Impact Factor\*** | **Total Citations** |
| --- | --- | --- | --- | --- |
| **1** | Pediatrics | 5 | 8.0 | 424 |
| **2** | Pediatric Infectious Disease Journal | 4 | 2.2 | 328 |
| **3** | BMC Infectious Diseases | 3 | 3.0 | 168 |
| **4** | Journal of Clinical Virology | 3 | 3.4 | 152 |
| **5** | Journal of Pediatrics | 2 | 4.4 | 536 |
| **6** | Journal of Clinical Microbiology | 2 | 6.1 | 299 |
| **7** | Other journals (n = 25) | 31 | — | 2,320 |

The geographic and organizational footprint is shown in Figure 4, revealing limited multicenter involvement. Only 2/50 papers (4%) were multicenter (one prospective, one retrospective cohort), while the remaining 48 (96%) are single-site or unspecified, raising concerns about external validity, especially where care pathways or laboratory methods vary by institution.

**Figure 4:**
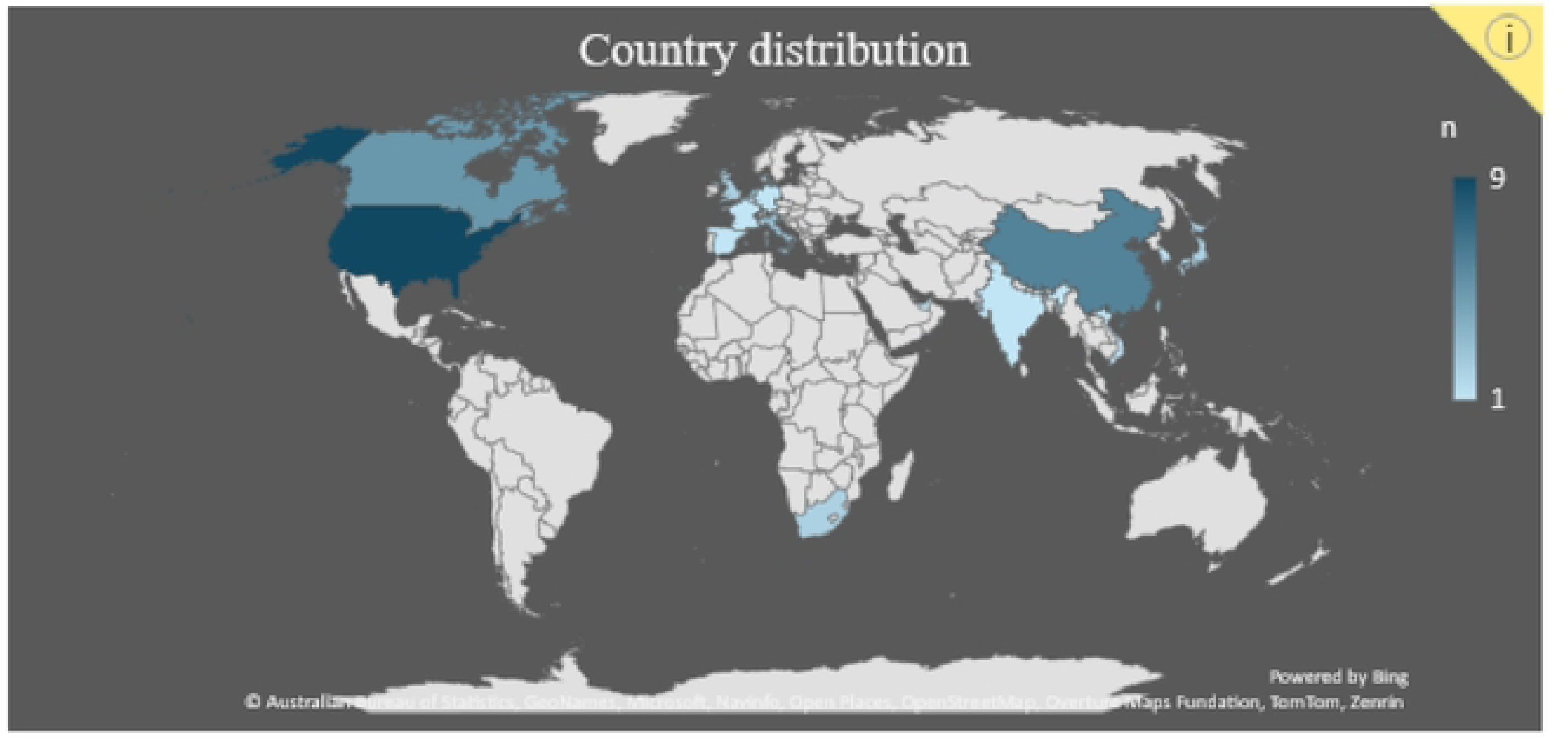
Article distribution by country.

The United States (9/50; 18%), China (6/50; 12%) and Canada (5/50; 10%) contributed most, followed by Taiwan, the Netherlands, and Italy (3 each; 6%), the United Kingdom, South Africa, and Japan (2 each; 4%), and single-paper contributions (2% each) include Germany, France, Denmark, Spain, South Korea, UAE, Vietnam, and Slovenia. Some regional aggregates (South America, West Africa, Latin America) were also recorded, potentially masking intra-regional variability. Overall, the evidence base is concentrated in North America, East Asia, and Western Europe and is predominantly single-centre, highlighting the need for broader geographic and multicenter participation to enhance generalizability and equity of evidence.

Stratification by evidence level, summarized in Table 5 and Figure 5, shows that 68% (34/50) are Level II (RCTs or well-designed comparative cohorts) and 16% (8/50) Level I syntheses, suggesting scope for more integrative reviews and meta-analyses. Lower hierarchy evidence includes Level IV case series/retrospective analyses (6%; 3/50), Level III non-randomized prospective studies (2%; 1/50), and Level V expert opinion or guideline articles (8%; 4/50).

**Figure 5:**
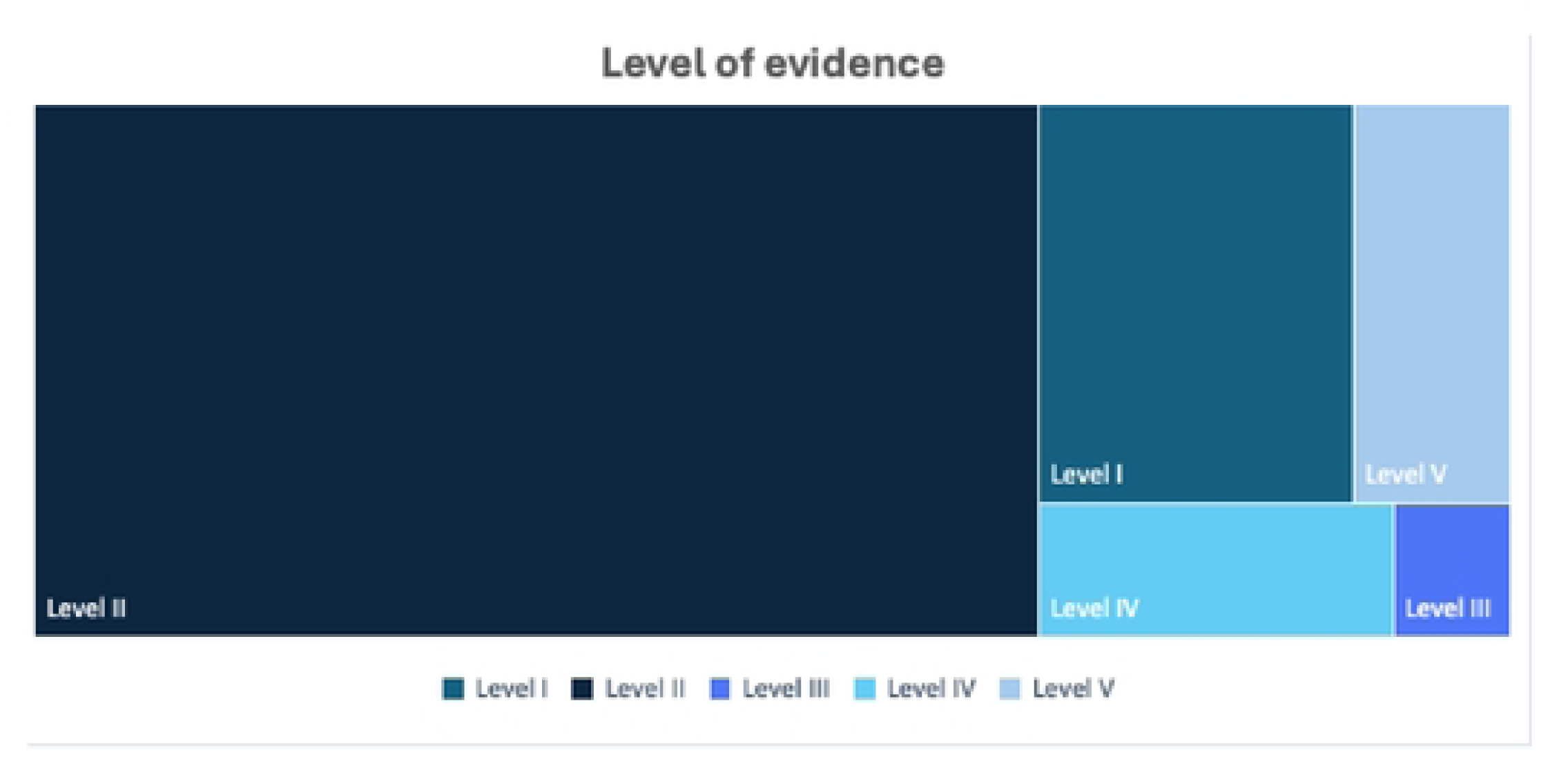
Level of evidence of the most-cited articles.

**Table 5:** Level of evidence of the most-cited articles.

| Level of Evidence | Number of Articles (n = 50) | % |
| --- | --- | --- |
| Level I – Systematic reviews / meta-analyses | 8 | 16.0 |
| Level II – RCTs or comparative studies with control | 34 | 68.0 |
| Level III – Non-randomized prospective studies | 1 | 2.0 |
| Level IV – Case series / retrospective analyses | 3 | 6.0 |
| Level V – Expert opinion / guideline papers | 4 | 8.0 |

Citation impact is driven by controlled comparative designs, with fewer top-tier syntheses than ideal for continuous guideline-level updating.

Observational studies dominate, cohort design account for two-thirds of the corpus - Prospective cohorts lead (36%), followed by retrospective cohorts (30%). Experimental evidence is limited (RCTs 12%). Formal syntheses are a minority (systematic review plus meta-analysis, 6%; systematic review, 2%), while Review articles and Cross-sectional studies contribute 8% and 6%, respectively. This emphasizes continued reliance on observational cohorts to study treatment effects and prognostic factors, with opportunities to expand RCTs and high-level syntheses to strengthen guideline development.

The co-citation map illustrated in Figure 6 exhibits a multi-clustered yet interconnected structure. A central cluster (red) is anchored by Jain et al., 2015 (NEJM) and Bradley et al., 2011 (Clin Infect Dis), reflecting high-impact work on pediatric community-acquired pneumonia etiology and clinical guidance. A second cluster (green) centers around Gonzales, 1999 (JAMA), Nyquist, 1998 (JAMA), and Hersh, 2013 (Pediatrics), representing antibiotic overuse and stewardship literature. A virology-discovery cluster (orange/purple) is organized around van den Hoogen, 2001 (Nat Med), Allander, 2005 (PNAS), Williams, 2004 (NEJM), and Jartti, 2004 (EID)—highlighting molecular identification of novel respiratory viruses (eg., hMPV, bocavirus) and diagnostic advances. The blue cluster contains clinical epidemiology and pediatric infectious disease studies (eg., Koskenvuo 2008; Chávez-Bueno 2007), mapping severity, risk factors, and inpatient outcomes. Peripheral nodes (eg., Ujiie, 2021; Foley, 2021; Li, 2022, Lancet) reflect pandemic-era surveillance and burden updates linking discovery and clinical management.

**Figure 6:**
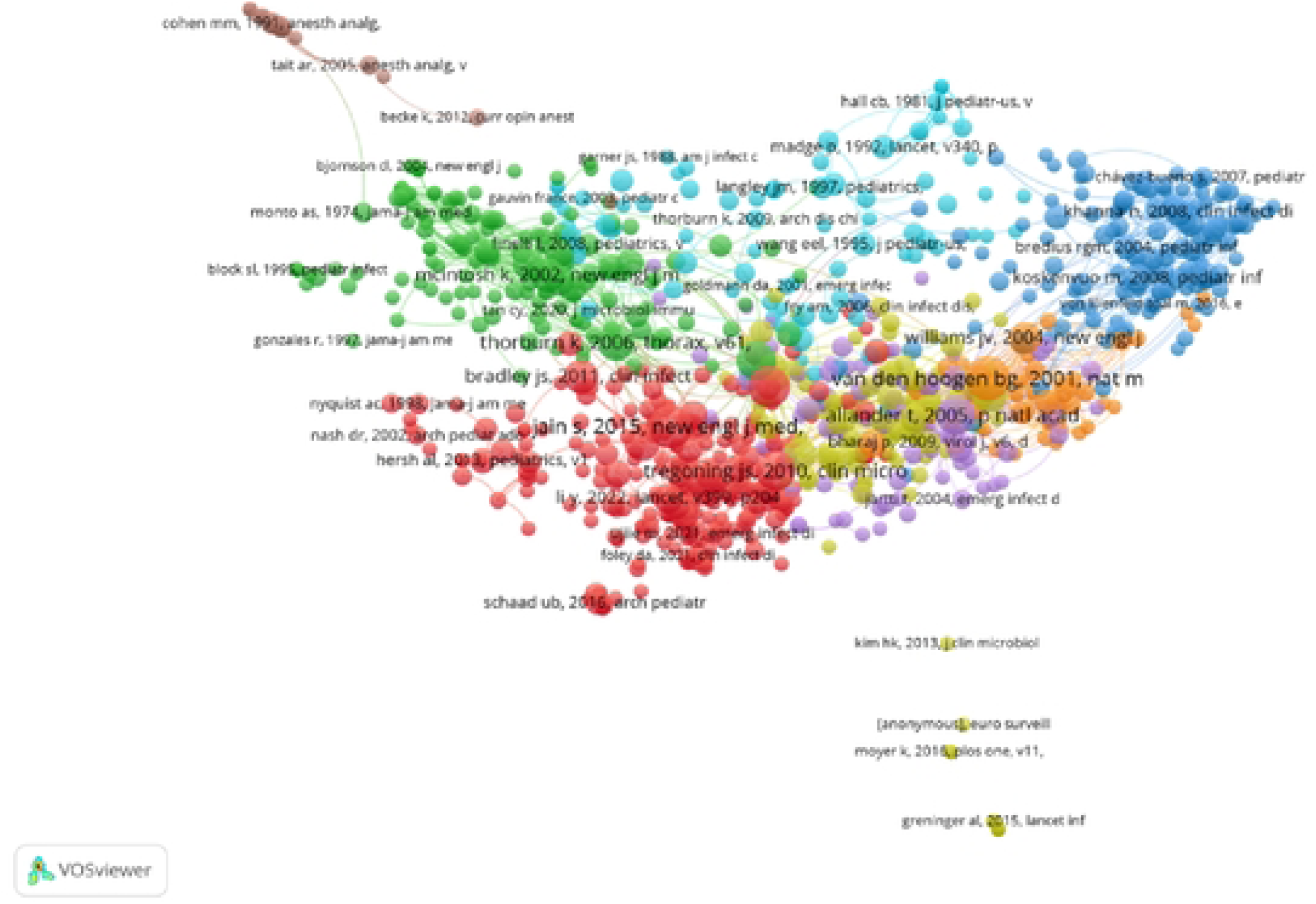
Co-citation map. The map illustrates multi-clustered, interconnected citation patterns. **Red cluster:** high-impact pneumonia etiology and clinical guidance studies. **Green cluster:** antibiotic overuse and stewardship literature. **Orange/purple cluster:** molecular discovery of novel respiratory viruses and diagnostic advances. **Blue cluster:** clinical epidemiology and pediatric infectious disease outcomes. Node size represents citation weight; dense intra-cluster links indicate self-reinforcing subfields, while sparser inter-cluster links highlight opportunities to integrate discovery with stewardship research.

Structurally, Jain 2015 and Bradley 2011 act as hub connecting stewardship and virology clusters, while van den Hoogen 2001 / Allander 2005 serve as foundational anchors for diagnostic/molecular threads that cross-cite clinical outcome studies. Node sizes (citation weight) and dense intra-cluster linkages suggest self-reinforcing subfields, whereas sparser links between stewardship and discovery indicate a need to integrate molecular diagnostics and pathogen-specific findings into practice-changing stewardship and outcome trials.

Overall, the map portrays a field whose citation influence is concentrated in landmark etiology, discovery, and guideline papers, with newer surveillance studies starting to link the clusters more closely.

The keyword co-occurrence map (Figure 7) highlights a dense, clinically oriented core anchored by “children,” “pediatric,” and “respiratory tract infection(s)”, which function as bridges linking several thematic clusters. One cluster concentrates on syndromes and outcomes (e.g., acute otitis media, pneumonia), while a second captures viral etiology and diagnostics (e.g., RSV, identification, transmission). A third cluster foregrounds antimicrobial therapy and stewardship (antimicrobial use, audit), with “antibiotic resistance” appearing peripherally, indicating a specialized, less interlinked niche within the highly cited set. A smaller, research-mechanism cluster centers on innate immunity/inflammation and environmental modifiers like air pollution, and a peri-procedural/critical-care edge (e.g., anesthesia, hypoxemia) is visible but loosely connected. The proximity of PCR/identification/RSV to pneumonia/acute otitis media suggests active translation from pathogen detection to disease-specific management, whereas sparser ties between the stewardship and immuno-inflammatory clusters highlight an integration gap—pointing to opportunities for trials linking rapid diagnostics and immune phenotyping to antibiotic decisions and patient-centred outcomes.

**Figure 7:**
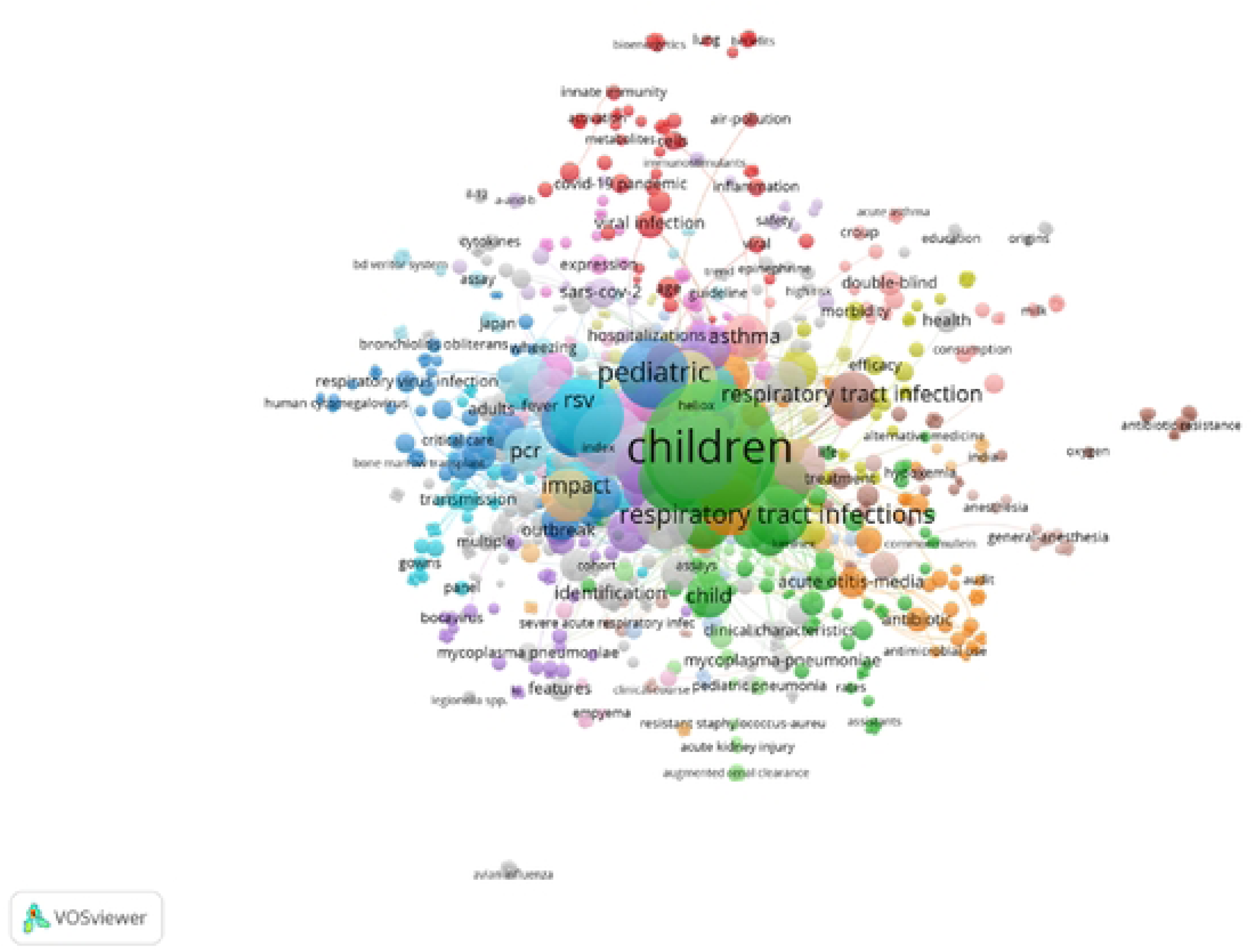
Keyword co-occurrence map. The map highlights a dense, clinically oriented core anchored by the terms “children,” “pediatric,” and “respiratory tract infection(s),” which link multiple thematic clusters. Key clusters include syndromes and outcomes (eg., acute otitis media, pneumonia), viral etiology and diagnostics (eg., RSV, identification, transmission), antimicrobial therapy and stewardship, and innate immunity/inflammation with environmental modifiers. Cluster proximity suggests translation from pathogen detection to disease-specific management, while sparser links between stewardship and immuno-inflammatory clusters indicate opportunities for integrating rapid diagnostics with patient-centered interventions.

## Discussion

This bibliometric study of the 50 most-cited pediatric respiratory infection articles shows progress in causation, management, and prevention, despite methodological and geographic gaps. Risk factors in developing countries include low birth weight, malnutrition, poverty, and low parental education [14–21].

Analyzing the 50 most-cited studies on pediatric respiratory infections reveals important patterns in research focus, study design, and global representation. The citation distribution is markedly right-skewed, with counts ranging from 384 at the top to 34 at the cutoff. A steep drop is observed after the top ten papers, showing that only a few landmark studies have exerted a major influence on the field, while most have received comparatively limited attention. The top ten papers hold a citation range of 384 to 110, reflecting their role as highly influential contributions. In contrast, the studies ranked 11 to 20 show a lower but still substantial citation range of 107 and 68. This pattern clearly distinguishes ‘landmark’ and ‘influential’ tiers within the literature. Additionally, a small number of highly cited studies—particularly large cohort investigations—were found to dominate scholarly attention in the field. The decade of the 2010s, when methodological consolidation occurred and pediatric respiratory disorders gained increased attention, saw the publication of over half of the most-cited studies. On the other hand, citation latency has prevented more recent contributions from the 2020s from gaining comparable importance, despite their promise. This is supported by bibliometric evidence, which indicates that the number of publications and citations in research on pediatric respiratory infections increased steadily between 2011 and 2022, reaching a peak of 133 and 136 publications in 2021 and 2022, respectively, and continuing to grow through 2023, reflecting the COVID-19 pandemic’s accelerating effects [22]. Although the number of publications has increased since 1980, a marked increase occurred from 2019 onwards, even though influenza and RSV cases decreased during the COVID-19 pandemic, with a notable resurgence of RSV infections in 2021 and late 2022, indicating that future research on infantile bronchiolitis is likely to see a new upturn in publications aimed at offering plausible explanations for this surge in cases [23].

An analysis of study design revealed that prospective and retrospective cohort studies, which have been crucial in shedding light on disease prevalence, risk factors, and outcomes, accounted for 66% of all research. Although they were present, randomized controlled trials (12%) and systematic syntheses (8%) are nevertheless relatively underrepresented, highlighting the field’s persistent dependence on observational data as its foundation.

Although extensive epidemiological mapping has been made easier by this focus, it has hindered the development of more robust causal conclusions and recommendations at the guideline level. Three main groups emerged from thematic mapping: (i) the epidemiology and clinical outcomes of infections, including bronchiolitis and pneumonia; (ii) the etiology and diagnostics of viruses; and (iii) the use and stewardship of antibiotics. Although these clusters were connected, there were integration gaps, especially between stewardship measures and rapid molecular diagnostics. This implies that systematic management systems have not yet fully incorporated diagnostic innovation.

Geographically, high-income nations like the United States, China, and Canada made the most significant contributions. Just 4% of the studies were multicenter in character, and there was very little representation from low- and middle-income nations. According to a bibliometric review of research on pediatric Mycoplasma pneumoniae, 86 countries and regions contributed, with China accounting for the largest share of publications (34.4%), followed by the US, Japan, Italy, and South Korea [24]. A persistent disparity in publication number and citation impact was highlighted by another bibliometric analysis, which found that China (67 studies, 3426 citations) and the United States (76 studies, 3751 citations) once again dominated, followed by Turkey, Italy, England, and France [25]. Similarly, RSV prevention papers were shown to originate from 90 countries, led by the United States (349 publications) [26]. Sparse data from low-resource settings, which carry a disproportionate pediatric respiratory infection burden, limits the external validity of these findings.

Journal representation revealed that citation influence was concentrated in a few high-profile publications, even though the 50 articles were spread over 31 journals. While journals like the Journal of Pediatrics earned high citation counts despite fewer publications, Pediatrics contributed the biggest percentage of papers. This highlights the critical role that specialized clinical platforms play in influencing visibility and practice. In parallel, bibliometric studies using Bradford’s Law have consistently shown that a limited set of journals drives a disproportionate share of scholarly influence [26][27]. One review of pediatric infectious disease research identified 12 core journals, the majority based in the United States, with Pediatrics achieving the highest citation rate and Vaccine, the Pediatric Infectious Disease Journal, and the Journal of Virology leading in publication volume [26].

When put together, the current bibliometric profile shows a developed but unbalanced field. While randomized trials, systematic syntheses, multicenter partnerships, and LMIC-driven research are still relatively rare, it is based on observational evidence and is dominated by high-income nations. To create strong, universally applicable, and fair guidelines for the care and prevention of pediatric respiratory infections, these deficiencies must be filled.

## Conclusion

Pediatric respiratory infections remain life-threatening worldwide. This study reviews the 50 most-cited papers, integrating methodology, research design, clinical practice, public health policy, and scientific contributions. Most research originates from high-income countries, with low- and middle-income nations underrepresented. Identified, influential studies rely largely on observational designs, while randomized trials, systematic reviews, and multicenter or low-income–country–led studies are uncommon.

## Data Availability

No data was generated by this study. The following existing data sources were used: [Top 50 Cited Articles on Pediatric Respiratory Infections] from [Web of Science].

## Acknowledgments

None.

